# Neurofeedback-guided kinesthetic motor imagery training in Parkinson’s disease: Randomized trial

**DOI:** 10.1101/2022.01.05.22268816

**Authors:** Sule Tinaz, Serageldin Kamel, Sai S. Aravala, Mohamed Elfil, Ahmed Bayoumi, Amar Patel, Dustin Scheinost, Rajita Sinha, Michelle Hampson

## Abstract

**Background:** Parkinson’s disease (PD) causes difficulty with maintaining the speed, size, and vigor of movements, especially when they are internally generated. We previously proposed that the insula is important in motivating intentional movement via its connections with the dorsomedial frontal cortex (dmFC). We demonstrated that subjects with PD can increase the right insula-dmFC functional connectivity using fMRI-based neurofeedback (NF) combined with kinesthetic motor imagery (MI). The current study is a randomized clinical trial testing whether NF-guided kinesthetic MI training can improve motor performance and increase task-based and resting-state right insula-dmFC functional connectivity in subjects with PD.

**Methods:** We assigned nondemented subjects with mild PD (Hoehn & Yahr stage ≤ 3) to the experimental kinesthetic MI with NF (MI-NF, n=22) and active control visual imagery (VI, n=22) groups. Only the MI-NF group received NF-guided MI training (10-12 runs). The NF signal was based on the right insula-dmFC functional connectivity strength. All subjects also practiced their respective imagery tasks at home daily for 4 weeks. Post-training changes in 1) task-based and resting-state right insula-dmFC functional connectivity were the imaging outcomes, and 2) MDS-UPDRS motor exam and motor function scores were the clinical outcomes.

**Results:** The MI-NF group did not show significant NF regulation and was not significantly different from the VI group in any of the imaging or clinical outcome measures. The MI-NF group reported subjective improvement in kinesthetic body awareness. There was significant and comparable improvement only in motor function scores in both groups. This improvement correlated with NF regulation of the right insula-dmFC functional connectivity only in the MI-NF group. Both groups showed specific training effects in whole-brain functional connectivity with distinct neural circuits supporting kinesthetic motor and visual imagery (exploratory outcome).

**Conclusions:** The functional connectivity-based NF regulation was unsuccessful in our cohort with mild PD. However, kinesthetic MI practice by itself or in combination with other imagery techniques is a promising tool in motor rehabilitation in PD.

**Highlights:**

- Parkinson’s disease (PD) causes difficulty with sustained motor performance
- Insula and dorsomedial frontal cortex (dmFC) are implicated in motivating movement
- Regulation of insula-dmFC functional connectivity with neurofeedback (NF) failed
- Motor imagery practice regardless of NF improved motor function and body awareness
- Motor imagery is a promising strategy for motor rehabilitation in PD

## 1. Introduction

Parkinson’s disease (PD) is a neurodegenerative disorder that causes significant motor and nonmotor disability. Loss of dopaminergic neurons in the substantia nigra *pars compacta* is the pathological hallmark of the disease (Braak et al., 2003). The impaired ability to sustain a steady motor performance is a major cause of morbidity in patients with PD. This is characterized by a rapid progressive decrement in the speed, amplitude, or force of movements during continuous tasks (e.g., walking, writing) (Chee et al., 2009, Kang et al., 2011, Ling et al., 2012). The standard pharmacological (e.g., dopaminergic medications) and surgical therapies (e.g., deep brain stimulation) are ineffective in improving the decrement (Baraduc et al., 2013, Bologna et al., 2018, Espay et al., 2011). It has been shown that the decrement is most pronounced when patients with PD have to internally generate movement, and improves when they are provided external cues for movement (Demirci et al., 1997, Morris et al., 2008, Tinaz et al., 2016). This suggests that there is a problem with the internal motivation of movement in patients with PD (Mazzoni et al., 2007). Neuroimaging studies typically implicate the dysfunction of motor cortical-basal ganglia circuits as the neural underpinning of the difficulty with internally-generated movement in patients with PD (Berardelli et al., 2001). The dorsomedial frontal cortex (dmFC) regions, including the supplementary motor area (SMA), pre-SMA, and cingulate motor areas, are involved in intentional motor control including movement initiation and maintenance. Neuroimaging studies have shown reduced activation in these cortical regions and the basal ganglia during internally-generated sequential movements in patients with PD (Catalan et al., 1999, Jahanshahi et al., 1995, Nakamura et al., 2001, Sabatini et al., 2000, Samuel et al., 1997, Yu et al., 2007). However, dysfunction in limbic brain circuits pertaining to the internal drive behind intentional movement may also play a role in PD. In a previous paper, we proposed that the standard cortical-basal ganglia circuit model of motor dysfunction in PD needs to be expanded to include the insula (Tinaz et al., 2018). Briefly, we argued that the insula, by processing a wide range of sensory signals arising from the body and integrating them with the emotional and motivational context, provides the impetus for intentional movement via its connections with the dmFC regions. Furthermore, we demonstrated in a proof-of-concept experiment that the functional connectivity between the right insula and dmFC can be strengthened with neurofeedback (NF)-guided kinesthetic motor imagery using functional MRI (fMRI) in subjects with PD (Tinaz et al., 2018).

In this study, we aimed to further test the findings of the proof-of-concept experiment in a randomized trial to investigate whether kinesthetic motor imagery training combined with fMRI-based NF using the right insula-dmFC functional connectivity can improve motor performance in patients with PD. We also examined whether this connectivity-based training changes the task-based and intrinsic whole-brain functional connectivity as has been demonstrated in other studies (Megumi et al., 2015, Yamashita et al., 2017).

Motor imagery refers to the mental rehearsal of motor acts without overt body movements. Imagined movements share commonalities with real movements including similar neural substrates, autonomic responses, and duration that have led to the notion of “functional equivalence” (Guillot et al., 2014). Motor imagery has been used to enhance athletic performance (Guillot and Collet, 2008) and in stroke rehabilitation (Di Rienzo et al., 2014), but rarely in a systematic way in motor rehabilitation in PD even though it is considered a valid compensatory strategy, for example, for gait impairment in people with PD (Tosserams et al., 2020). Several studies using short-term motor imagery practice in PD cohorts have failed to demonstrate significant motor facilitation (Abraham et al., 2021, Caligiore et al., 2017).

However, in longer-term motor rehabilitation studies, it has been shown that patients with PD have the capacity to perform motor imagery of everyday actions to improve slowness (Tamir et al., 2007) and can increase motor imagery-related brain activation using fMRI-based NF (Subramanian et al., 2016, Tinaz et al., 2018).

Different types of motor imagery (e.g., kinesthetic vs visual) recruit different brain circuits (e.g., sensorimotor vs. visuospatial) (Guillot et al., 2009, Guillot et al., 2014, Lorey et al., 2009). Here, we specifically chose kinesthetic motor imagery training as the experimental intervention because this type of imagery preferentially recruits sensorimotor brain regions including the insula (Lorey et al., 2009, Marchesotti et al., 2016). The imagery training focused on gross motor tasks that belonged to the individual’s motor repertoire and activities of daily living (e.g., walking, gym exercises, shoveling snow). We used visual imagery training without NF as the active control condition. We hypothesized that kinesthetic motor imagery training with NF, but not visual imagery, would increase the task-based and resting-state right insula-dmFC functional connectivity strength and improve motor performance in patients with PD.

## 2. Methods

### 2.1. Overview

This study was designed as a phase 1 clinical trial (NCT03623386). Our design is in line with the recommendations in the “consensus on the reporting and experimental design of clinical and cognitive-behavioral neurofeedback studies (CRED-nf)” checklist (Ros et al., 2020). The sample size was determined based on moderate clinically meaningful improvement criteria as determined by the part III motor exam scores of the Movement Disorders Society-Unified Parkinson’s Disease Rating Scale (MDS-UPDRS) (Goetz et al., 2008). Two groups of subjects with PD were randomly assigned in parallel. The experimental group (MI-NF) received NF-guided kinesthetic motor imagery training and the active control group (VI) received visual imagery training without NF. The control group was not aware of the NF component in the experimental arm. Subjects in both groups were provided with explicit imagery strategies and practiced their respective imagery tasks daily for about four weeks. The primary clinical outcome assessor (A.P.) was masked to the intervention assignment.

Our choice of the control condition warrants further explanation: Our preliminary findings clearly suggested that frustration and mental disengagement during the task consistently generated negative NF in subjects with PD (Tinaz et al., 2018). Sham (i.e., noncontingent) NF is a commonly used control condition in NF paradigms. However, it can induce frustration and disengagement (Sorger et al., 2019, Stoeckel et al., 2014), hence, generate negative NF which might lead to overestimation of the effects of contingent NF. Therefore, we chose an active control condition instead of using sham NF to minimize this risk. We used visual imagery as the control task to account for the cognitive and attentional aspects of the experimental kinesthetic motor imagery task. This design also allowed us to match the duration of imagery practice between the groups throughout the study, and to facilitate motivation and active participation of the control group.

We used a bar graph representation of the functional connectivity strength between the right insula and dmFC as the NF modality. The measure of successful NF learning was the increase in this functional connectivity strength from the first (baseline) to the last (post-training) imagery scans that were performed without NF by all subjects. We used intermittent NF which has the following advantages over continuous NF: 1) avoiding potentially distracting confound of cognitive load associated with constant NF monitoring (Stoeckel et al., 2014), and 2) separating out brain activity related to NF evaluation and actual task performance (Johnson et al., 2012).

### 2.2. Subjects

We recruited subjects with PD defined according to the UK Brain Bank diagnostic criteria (Hughes et al., 1992) through the Yale Movement Disorders Clinic and via local PD support groups in Connecticut. All subjects participated in the study after giving written informed consent in accordance with the procedures approved by the Human Research Protection Office of the Yale School of Medicine. We conducted the study at the Yale Magnetic Resonance Research Center. Subjects underwent screening for MRI safety and were asked detailed questions about their clinical history to determine eligibility. Subjects with PD who were not fully independent, had a neurological or psychiatric disorder (other than PD and comorbid depression or anxiety), or a medical condition that might affect the central nervous system, history of alcohol or illicit drug abuse, head injury resulting in loss of consciousness, dementia (Montreal Cognitive Assessment (MoCA) score < 21), or contraindications for MRI were excluded (see Supplementary Material for details).

We enrolled the subjects continuously and randomized them to two groups using the “minimization” method (Zhao et al., 2015). After truly randomly allocating the first ten subjects, each subsequent subject was allocated to either group considering the factors age, gender, and disease stage. This method allowed us to minimize the imbalance between the groups in terms of demographic and clinical characteristics.

### 2.3. Sample size calculation

The sample size calculation was based on the clinically important difference (CID) in the primary motor outcome measure. Approximately a 5-point decrease in the mean MDS-UPDRS motor exam score after an intervention has been defined as a moderate CID (Schrag et al., 2006, Shulman et al., 2010). In this study, we expected a moderate CID. We used the mean and standard deviation of the MDS-UPDRS part III motor exam scores (24 ± 9) of a large group of subjects with PD (N = 347) as the population mean and standard deviation (Shulman et al., 2010). Assuming α = 0.05 and power = 0.80, 22 subjects were required for a moderate CID. Considering the possibility of a 15% drop-out rate, we planned to recruit 25 subjects for each group, a total of 50 subjects. We recruited 54 subjects (i.e., the exclusion rate was 18.5%). A total of 10 subjects were excluded: Two dropped out, one could not complete the visits due to Covid-related restrictions, one felt claustrophobic during the first MRI scan, one was found to have a cavernoma in the occipital cortex, and five (two in the VI group and three in the MI-NF group) were excluded due to homework noncompliance (i.e., < 50% completion rate of daily homework).

### 2.4. Clinical and psychometric evaluations

Subjects underwent all motor assessments in the medication “off” state (12-hr washout) because the “off” state reflects the disease severity more accurately. We assessed disease severity and stage using the MDS-UPDRS (Goetz et al., 2008) and the Hoehn and Yahr (H & Y) scale (Hoehn and Yahr, 1967). We videotaped the part III motor exam of the MDS-UPDRS for all subjects. A movement disorders neurologist (A.P.), who was masked to the group assignment and visit order of the subjects, viewed the videos and scored the exam features (except for rigidity, which was scored by S.T. during the actual exam).

The motor function tests included the two-minute endurance walking, timed up-and-go, five times sit-to-stand, 360-degree turning, and Physical Performance Test to measure the speed of gross movements. The Physical Performance Test measures the speed of everyday motor tasks (e.g., writing, simulated eating, putting on and removing a coat, climbing up the stairs, etc.) (Reuben et al., 1990).

To rule out dementia we administered the MoCA test (Nasreddine et al., 2005). In line with the MDS Task Force recommendations, we administered the Spielberger State-Trait Anxiety Inventory (STAI-S and -T) (Spielberger et al., 1983), Beck Depression Inventory-II (BDI-II) (Beck et al., 1996), and Starkstein Apathy Scale (Starkstein et al., 1992) to evaluate subjects for anxiety, depression, and apathy, respectively. In addition, we administered the Parkinson’s Fatigue Scale (PFS-16) (Brown et al., 2005) and PD Quality of Life Questionnaire (PDQ39) (Jenkinson et al., 1997). We used the 12-item Movement Imagery Questionnaire-3 (MIQ-3) (Williams et al., 2012) to assess subjects’ ability for visual and kinesthetic imagery of movements. For the VI group, we used the Scales for Outcomes of PD-cognition (Marinus et al., 2003) and additional surveys (proactive coping inventory (Greenglass & Schwarzer, 1998); self-efficacy (Chen et al., 2001); self-regulation (Schwarzer et al., 1999)) as “fillers” in visit 3.

### 2.5. MRI Scanning

We scanned the subjects in the morning or early afternoon after they took their dopaminergic medication. We collected the scans of 32 subjects in a 3.0 Tesla Siemens Trio TIM, and those of 12 subjects in a 3.0 Tesla Siemens Prisma scanner using a 32-channel head coil and identical sequences in both scanners. First, we collected high-resolution T1-weighted MPRAGE images (176 slices, voxel size: 1 mm^3^, FoV: 250 mm, matrix: 256 × 256, TR: 1900 ms, TE: 2.52 ms, TI: 900 ms, flip angle: 9 degrees) for an accurate localization of the functional MRI data in the beginning of each scan session. Then, we collected T1-weighted FLASH axial images (36 slices, voxel size: 0.9 × 0.9 × 4 mm, FoV: 224 mm, matrix: 256 × 256, TR: 300 ms, TE: 2.47 ms, flip angle: 60 degrees) as an intermediate scan to coregister MPRAGE and echo-planar functional images for the imagery scans. Finally, we obtained axial oblique T2*-weighted, echo-planar functional images (36 slices, voxel size: 3.5 × 3.5 × 4 mm, FoV: 224 mm, matrix: 64×64, TR: 2000 ms, TE: 25 ms, flip angle: 90 degrees). The number of acquisitions was 120 (4 min) for all imagery scans and 304 (10 min 8 s) for the resting-state scans. We always collected the resting-state scans first, during which subjects were instructed to keep their eyes closed, avoid any voluntary movement, and let their mind wander. We assessed wakefulness at the end of the scan by subjects’ report.

### 2.6. Study Protocol

The study protocol is summarized in Fig. 1. After screening, subjects were scheduled for an initial evaluation visit. Visit 2 was scheduled within five weeks after the initial visit. The subsequent visits were on average two weeks apart.

**Fig. 1.**
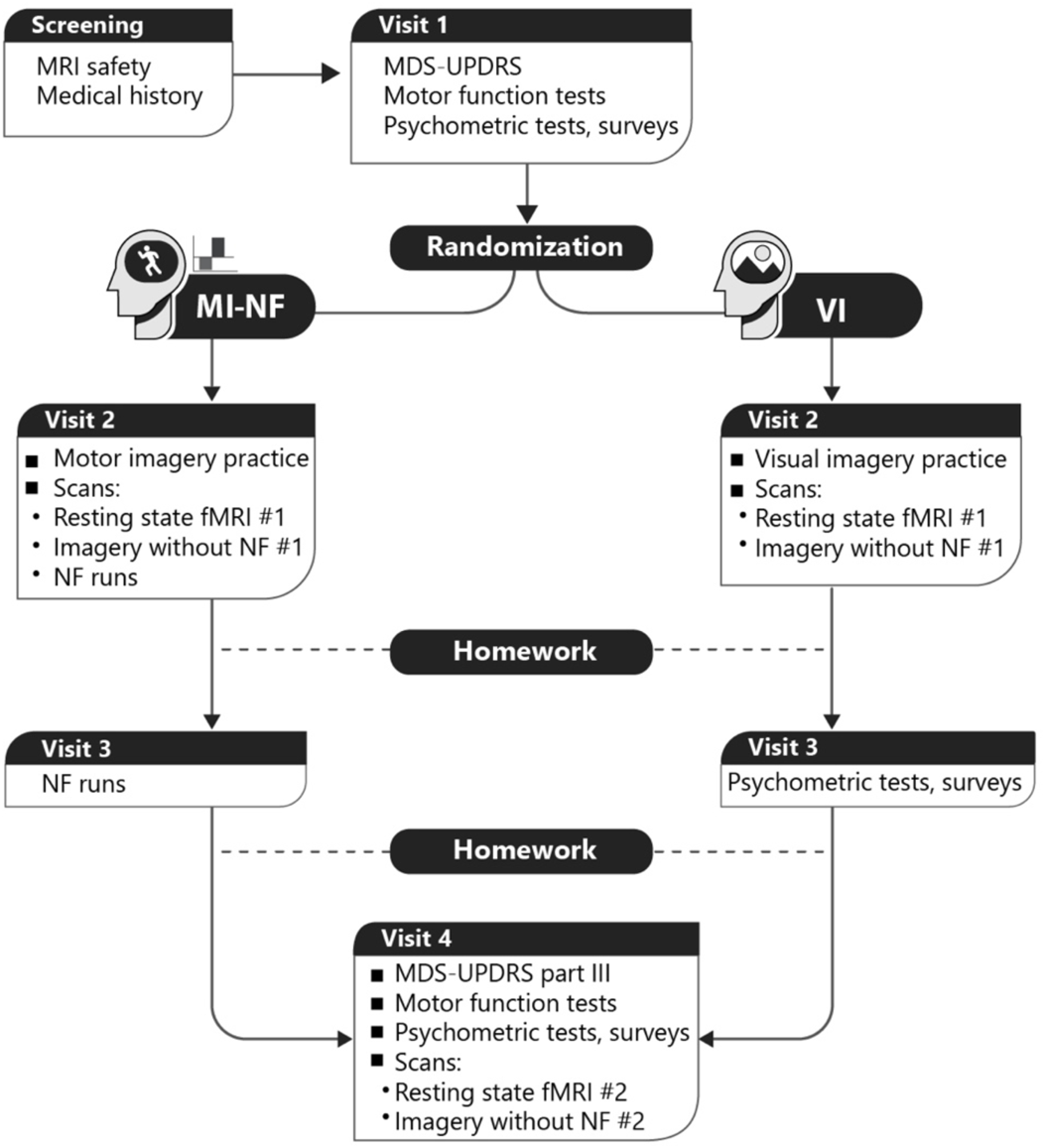
Study Protocol.

**Visit 1:** We performed the initial clinical and psychometric evaluations including the administration of the MDS-UPDRS, motor function tests, MoCA, BDI-II, STAI, Apathy Scale, PFS-16, PDQ39, and MIQ-3 in this visit. Upon completion of visit 1, we randomized the eligible subjects to either the VI or MI-NF group.

**Visit 2**: First, we reviewed the basic concepts and purpose of the respective imagery tasks with all subjects. We discussed the potential benefits of imagery practice in improving motor functioning and visuospatial attention with the MI-NF and VI groups, respectively, and emphasized the importance of consistent practice to achieve these benefits. We also provided the respective imagery strategies in written form.

*MI-NF group:* We determined each subject’s motor repertoire, identified their motor difficulties, and familiarized them with the kinesthetic motor imagery tasks. Subjects were primed for body awareness by engaging in a mindfulness body scan practice during which they listened to an audio recording guiding them to pay attention to sensations in different body parts. Subsequently, subjects practiced kinesthetic motor imagery of basic movements via audio-recorded scripts. Subjects first performed the movements (e.g., raise your knee, lift up your arm, tap your foot), then imagined performing the movements. Finally, subjects practiced kinesthetic motor imagery of complex whole-body movements of common activities (e.g., walking, balance exercises, calisthenics) via audio-recorded scripts and were instructed to focus on body sensations and positively-valenced emotions associated with imagined movements.

*VI group*: Subjects received guided visual imagery training via audio-recorded scripts. Subjects practiced visual imagery of static objects or scenery (e.g., tree, lake, sunset, mountain, special place) from a motionless first-person view focusing on sensory features such as colors, shapes, smells, sounds. There was no reference to body movements or sensations in the scripts and subjects were instructed to avoid movement in their imagery. Subjects were encouraged to evoke memories of familiar places or scenes during imagery and focus on positively-valenced emotions associated with the visual imagery content.

The MRI scans followed the practice session. All subjects completed the resting-state fMRI scan first. Subsequently, all subjects practiced their respective baseline imagery task in the scanner during which no NF was provided. Subjects in the MI-NF group then started the NF-guided kinesthetic motor imagery sessions in the scanner. They were free to choose the content of imagery, but told not to change it within a block. There were 4-5 NF training runs each lasting 4 min for the MI-NF group in Visit 2. The VI group did not receive any NF training sessions. All subjects were debriefed about their experience and imagery content after scanning. The MI-NF group was encouraged to use the “winning” strategies in NF sessions during homework practice.

*Homework*: Upon completion of scanning, we instructed subjects on how to perform and report their daily imagery homework using the Yale Qualtrics online survey platform. The MI-NF group was instructed to practice mindfulness body scan and kinesthetic motor imagery via audio-recorded scripts at home every day for two weeks until Visit 3. We structured the online homework survey based on the key components of motor imagery practice including the setting, type of activity, vividness, difficulty, duration, as well as the type and quality of body sensations and emotions evoked by the imagined movements (Collins and Carson, 2017). Subjects in the VI group were also instructed to continue practicing visual imagery exercises via audio-recorded scripts at home every day for two weeks until Visit 3. The online homework survey included questions about the setting, content, sensory modalities, vividness, difficulty, and duration of the visual imagery practice, as well as the emotions evoked by this practice. All subjects responded to the survey questions by selecting existing options for each respective imagery component and also entered free text. All subjects received daily reminders via emails to ensure adherence to daily imagery exercises.

**Visit 3**: First, we reviewed the homework of all subjects and discussed strategies for improvement and refinement of their imagery practice as needed. Then, the MI-NF group completed another set of 6-7 NF training runs in Visit 3. The VI group completed psychometric tests and surveys, which served as “fillers” to match the exposure time.

Both groups continued their respective imagery practices at home and completed the online homework surveys until Visit 4.

**Visit 4**: Subjects held the morning dose of their dopaminergic medications. The MDS-UPDRS part III motor exam, motor function tests, and MIQ-3 were repeated on all subjects. We discussed with all subjects in the MI-NF group whether they noticed any improvement in the specific motor difficulties that they reported in the first visit. After these assessments, subjects took their medication and completed a resting-state scan as well as an imagery scan during which they practiced kinesthetic motor and visual imagery, respectively, without NF.

### 2.7. Outcome measures

The primary imaging outcome measure was the change in right insula-dmFC functional connectivity strength 1) from the first to the last respective imagery scans without NF and 2) from the first to the last resting-state scans. We hypothesized that the MI-NF group would show a significant increase in both functional connectivity measures compared with the VI group.

The primary clinical outcome measure was the change in MDS-UPDRS part III motor exam scores from the first to the last visits. The secondary clinical outcome measure was the change in motor function test scores from the first to the last visits. We hypothesized that the MI-NF group would show a significant improvement in both clinical measures compared with the VI group.

We also examined the relationship between the imaging and clinical outcome measures in each group. We hypothesized that the increase in the right insula-dmFC functional connectivity would correlate with improvement in MDS-UDRS part III and motor function test scores only in the MI-NF group.

In addition, change in MIQ-3 scores and change in whole-brain resting-state and imagery task-based functional connectivity without NF were exploratory outcome measures. We hypothesized improvement in MIQ-3 scores in both groups and imagery task-specific changes in functional connectivity in both groups in the motor and visual networks, respectively.

### 2.8. Clinical and psychometric data analysis

We first assessed the normality of distribution of all clinical scores using the Shapiro-Wilk test. We compared the means and standard deviations of the normally distributed, and medians and median absolute deviations of the non-normally distributed scores with the population means or cutoff scores of the respective tests, when applicable, using one-sample t-tests (p < 0.05, two-tailed). For between-group comparisons, we used the two-sample t-test for normally distributed and nonparametric Mann-Whitney U test for non-normally distributed continuous variables (p < 0.05, two-tailed). We used the Chi-Square test for a between-group comparison of the categorical variables including handedness and disease onset side (p < 0.05, two-tailed). We used repeated measures ANOVA (within-subject: time with two levels, i.e., pre- and post-training; between-subject: group) to evaluate the between-group changes in the MDS-UPDRS part III exam scores, motor function test performances, and MIQ-3 scores as a result of training (p < 0.05, two-tailed). We used the SPSS 26 software for all statistical analyses.

### 2.9. Homework analysis

The number of entries and total time investment in homework, vividness, and difficulty of imagery were numerical values and averaged. Vividness and difficulty of imagery were rated on a Likert-like scale from 0 to 10, 0: not vivid/not difficult and 10: very vivid/very difficult. We compiled the selected and free-text responses in each imagery component under categories and calculated the percentages.

### 2.10. Imaging Data Analysis

#### 2.10.1. Right insula – dmFC functional connectivity during motor imagery scans with NF

A subdued background image of a night sky instructed subjects to engage in kinesthetic motor imagery for 40-s blocks (total of five blocks). At the end of each kinesthetic motor imagery block, NF was plotted as a bar graph (blue: negative, red: positive feedback) and presented for 8 s. NF was defined as the functional connectivity between the right insula and dmFC. We created cubic anatomical masks (6 × 6 × 6 mm) centered at x = 44, y = 4, z = 8 in the right insula and at x = −4, y = 2, z = 62 in the dmFC (Fig. 2). These regions of interest were functionally determined using a silent heartbeat counting task as described before (Tinaz et al, 2018). These masks were created based on the standard Montreal Neurological Institute (MNI) brain template and then registered to each subject’s native functional space using a series of transformations. All transformations were estimated using the Bioimage Suite software and manually inspected for accuracy. Functional scans of each subject were motion-corrected in real-time using the algorithms described in Scheinost et al. (2013). Covariates of no interest were regressed out including linear and quadratic drifts; mean cerebral spinal fluid, white matter, and gray matter signals; and motion-related confounds (24-parameter motion model including six rigid-body motion parameters, six temporal derivatives, and these terms squared). The data were temporally smoothed with a zero-mean unit variance Gaussian filter (approximate cutoff frequency = 0.12 Hz). Then, the signal time course of the right insula and dmFC masks in a given subject were computed as the average time course across all voxels within each of these masks. Finally, the right insula-dmFC functional connectivity was obtained by correlating the time courses and Fisher z-transforming the r-values (Scheinost et al., 2020). A Matlab program plotted the *z*-values as a bar graph, which was presented to the subjects in the MI-NF group as NF.

**Fig. 2.**
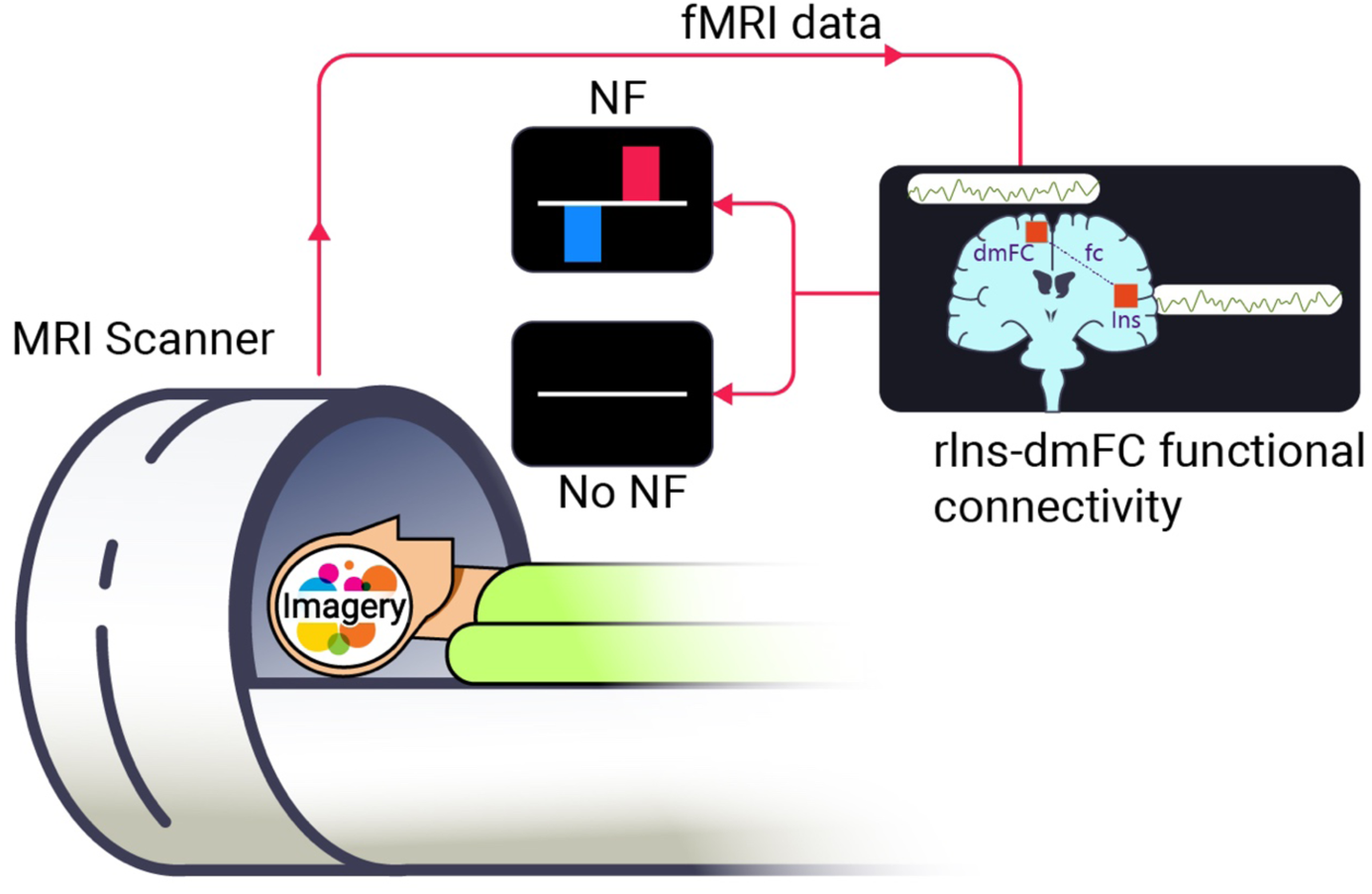
Neurofeedback brain-computer interface. Subjects in the MI-NF group performed motor imagery in the scanner during task blocks and received NF in the form of red (positive) and blue (negative) bars. The height of the bars reflected the strength of the positive/negative right insula-dmFC functional connectivity (fc). During the first and last imagery sessions without NF, all subjects performed their respective imagery in the scanner during task blocks and received no NF (flat line). Subjects in the VI group never received NF.

#### 2.10.2. Right insula – dmFC functional connectivity during motor and visual imagery scans without NF

We used the same task design and followed the same steps to compute the right insula-dmFC functional connectivity during imagery scans without NF as described in section 2.10.1. However, instead of bar graphs representing the z-values, a flat line (i.e., no NF) was presented. Subjects were explicitly told not to expect NF during these scans.

We calculated the percent change in the right insula-dmFC functional connectivity from the first to the last respective imagery scans without NF and performed a between-group comparison using the nonparametric Mann Whitney U test (p < 0.05, two-tailed).

#### 2.10.3. Analysis of the resting-state fMRI scans

We used the Connectivity toolbox v17 for the resting-state data analysis (Whitfield-Gabrieli and Nieto-Castanon, 2012). Preprocessing steps included the removal of the first four scans to reach magnetization steady state, motion correction, outlier detection (frame-wise displacement above 0.9 mm or global signal changes above 5 standard deviations), coregistration of functional scans with the structural scan, normalization to the standard MNI brain template, and smoothing with a Gaussian kernel with a FWHM of 8 mm to account for inter-individual anatomical variability. De-noising steps included correction for physiological and other sources of noise by regressing out the principal components of the white matter and cerebrospinal fluid signal using the CompCor method (Chai et al., 2012), regression of motion artifacts and outliers before bandpass-filtering, and linear detrending. Global signal was not removed. Finally, we bandpass-filtered (0.008 < f < 0.09 Hz) the data to capture the fluctuations of the blood oxygenation level-dependent (BOLD) signal that typically occur within this frequency range at rest.

For the whole-brain functional connectivity analyses, we used the functionally defined nodes (n = 268) of the whole-brain Shen atlas (Shen et al., 2013). For each subject, we extracted the average BOLD signal time courses from these nodes and correlated them with each other using Pearson correlations. The r-values corresponded to the functional connectivity strength between node pairs. We Fisher z-transformed the r-values and obtained group-level functional connectivity maps for statistical analyses. We used a 2 × 2 mixed ANOVA (within-subject: time with two levels, i.e., pre- and post-training; between-subject: group) for a between-group comparison of the training effect. We used the false discovery rate (FDR) method for correction for multiple comparisons (p < 0.05, two-tailed) (Genovese et al., 2002).

For the right insula-dmFC functional connectivity analyses, we extracted and correlated the resting-state BOLD signal time courses from these regions as explained in the previous paragraph. We then calculated the percent change in the right insula-dmFC functional connectivity from the first to the last resting-state scan and performed a between-group comparison using the nonparametric Mann Whitney U test (p < 0.05, two-tailed).

#### 2.10.4. Whole-brain connectivity analyses of imagery scans without NF

We applied the same preprocessing and denoising steps to the imagery scans as explained in section 2.10.3 with the following differences: The first three scans were removed and the BOLD signal was high-pass filtered [0.008 < f < Inf]. We then used the generalized psychophysiological interaction (gPPI) model in the Connectivity toolbox v17 (Whitfield-Gabrieli and Nieto-Castanon, 2012) to assess the imagery task-based whole-brain functional connectivity changes in each group. We convolved the imagery blocks and feedback periods separately with the canonical hemodynamic response function. We used the same functional connectivity calculations based on the imagery blocks and the 2 × 2 mixed ANOVA (within-subject: time with two levels, i.e., pre- and post-training; between-subject: group) as explained in section 2.10.3 for between-group comparison of the training effect (FDR-corrected p < 0.05, two-tailed).

Of note, to address the potential confounding effects of different scanners, we compared the voxel-wise global mean correlations (GCOR) for resting-state and imagery scans after denoising between subjects scanned in the Trio TIM and Prisma scanners.

#### 2.10.5. Correlations between imaging and clinical outcomes

We calculated the average percent change across all motor function test scores and MDS-UPDRS part III scores for each group. We then performed Kendall’s tau-b nonparametric bivariate correlations with bootstrapping (x1000) between the average percent change in motor function and part III scores and percent change in the 1) imagery task-based and 2) resting-state right insula-dmFC functional connectivity separately for each group (p < 0.05, two-tailed).

## 3. Results

### 3.1. Clinical and psychometric data at baseline

There was no significant difference between the MI-NF and VI groups in any of the demographic, clinical, and psychometric variables listed in Table 1. Moreover, both groups scored significantly below the anxiety, apathy, minimal depression, and fatigue cutoff scores; significantly above the MoCA cutoff score for mild cognitive impairment, and had a significantly better quality of life compared with the PD population with the same disease stage of H & Y 2 (see Supplementary Material Section 2 and Tables S1 and S2). The details of the medication types and LEDD distribution are shown in Tables S3 and S4 in the Supplementary Material.

**Table 1.**
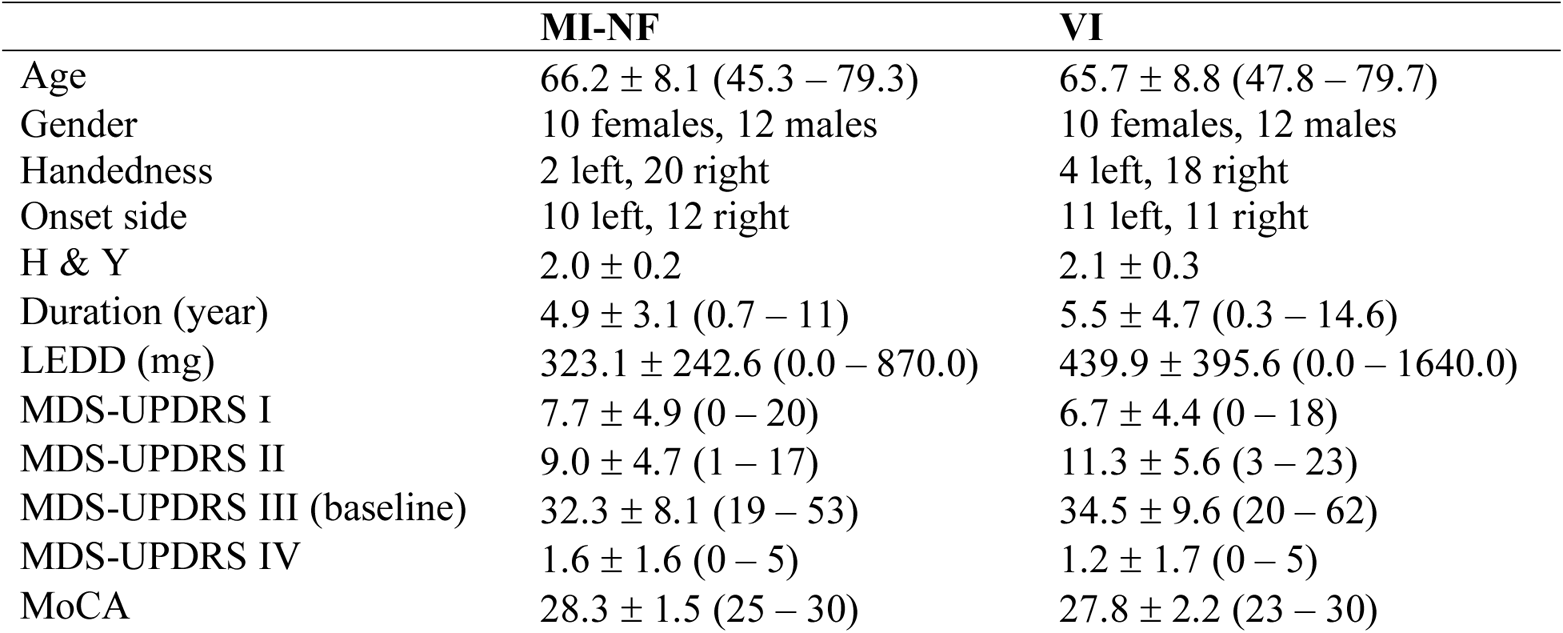
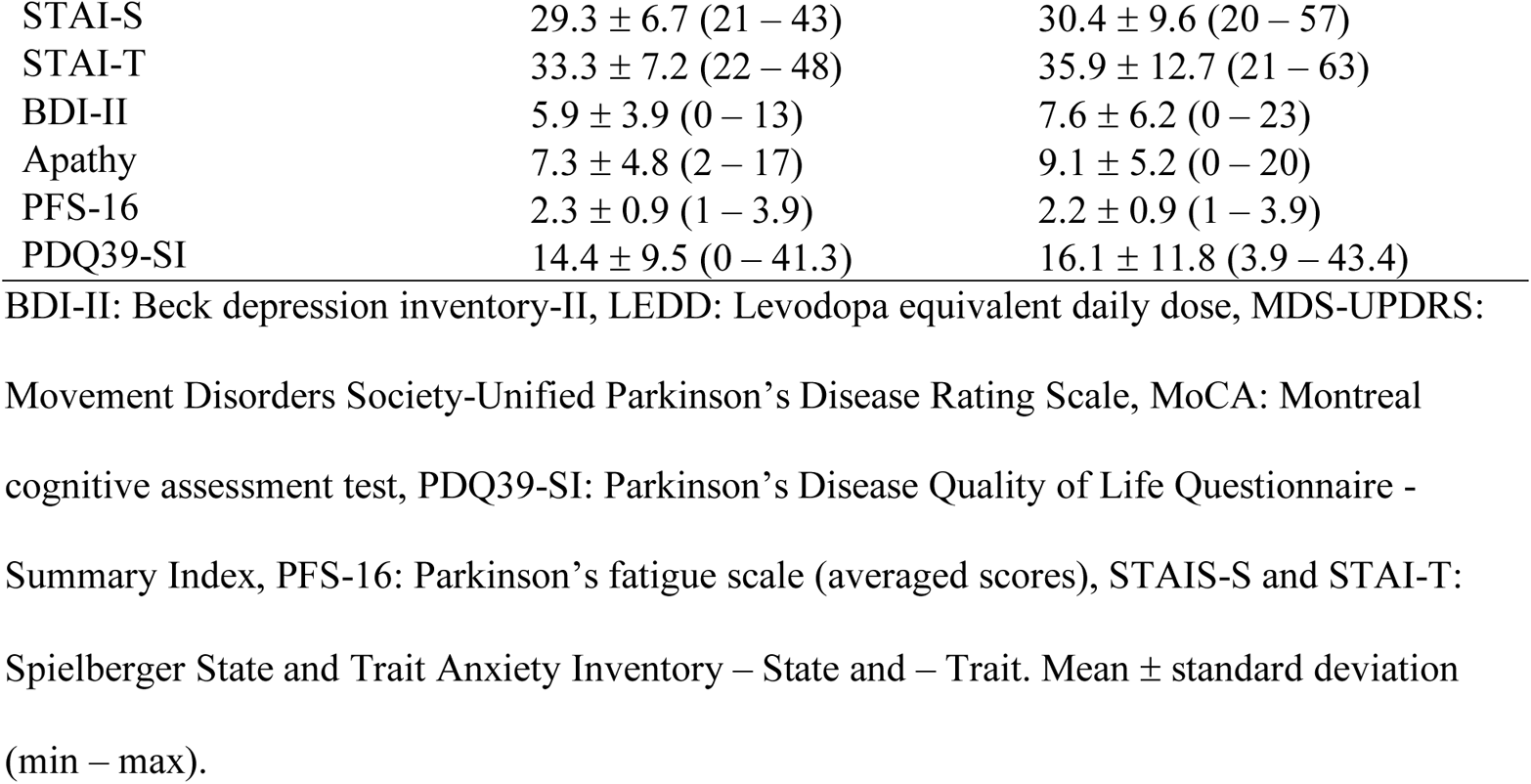
Clinical and psychometric data.

### 3.2. Training effects on behavior

The repeated measures ANOVA revealed no significant main effect of group, no significant time × group interaction, but significant main effect of time indicating that both groups significantly improved their gross motor performance and motor imagery skills post-training (Physical Performance Test: F(1,42) = 8.967, p = 0.005, endurance walking: F(1,42) = 18.276, p = 0.000, gross motor combined: F(1,42) = 5.309, p = 0.026; MIQ-3: F(1,42) = 6.553, p = 0.014).

However, the repeated measures ANOVA on the MDS-UPDRS part III scores did not show any significant main effect of group (F(1,42) = 1.204, p = 0.279), time F(1,42) = 0.072, p = 0.790), or significant time × group interaction (F(1,42) = 1.085, p = 0.303) (Table 2). A power analysis based on the mean difference and standard deviation between the pre- and post-training MDS-UPDRS part III scores of the MI-NF group revealed an effect size of 0.169 (α = 0.05 and power = 0.118). In other words, for α = 0.05 and power = 0.80, 276 subjects would be needed to see a significant change in the part III scores.

**Table 2.**
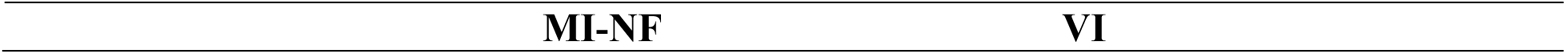
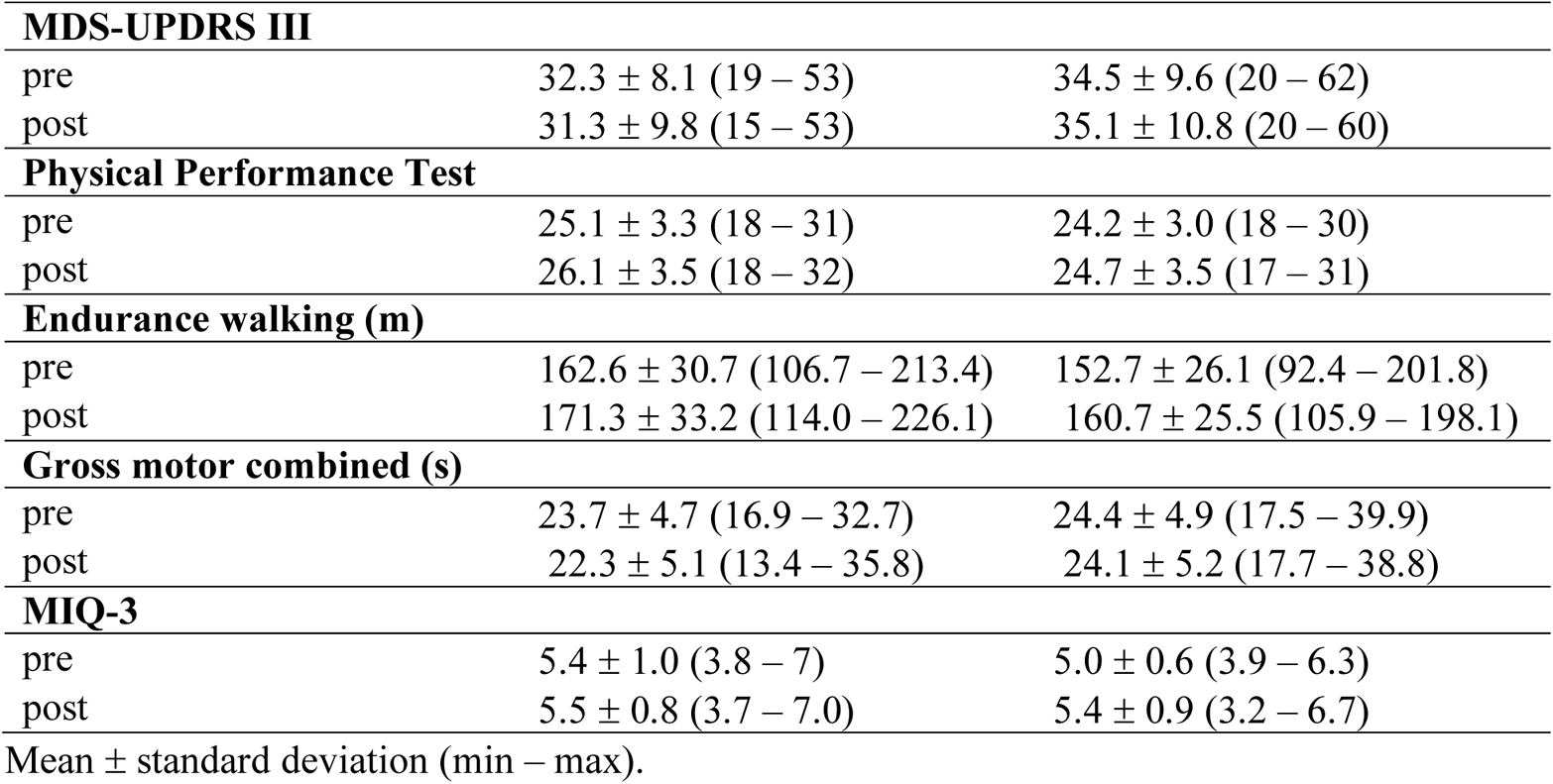
Pre- and post-training behavioral data.

|  | MI-NF | VI |
| --- | --- | --- |
| **MDS-UPDRS III** |  |  |
| pre | 32.3 ± 8.1 (19 – 53) | 34.5 ± 9.6 (20 – 62) |
| post | 31.3 ± 9.8 (15 – 53) | 35.1 ± 10.8 (20 – 60) |
| **Physical Performance Test** |  |  |
| pre | 25.1 ± 3.3 (18 – 31) | 24.2 ± 3.0 (18 – 30) |
| post | 26.1 ± 3.5 (18 – 32) | 24.7 ± 3.5 (17 – 31) |
| **Endurance walking (m)** |  |  |
| pre | 162.6 ± 30.7 (106.7 – 213.4) | 152.7 ± 26.1 (92.4 – 201.8) |
| post | 171.3 ± 33.2 (114.0 – 226.1) | 160.7 ± 25.5 (105.9 – 198.1) |
| **Gross motor combined (s)** |  |  |
| pre | 23.7 ± 4.7 (16.9 – 32.7) | 24.4 ± 4.9 (17.5 – 39.9) |
| post | 22.3 ± 5.1 (13.4 – 35.8) | 24.1 ± 5.2 (17.7 – 38.8) |
| **MIQ-3** |  |  |
| pre | 5.4 ± 1.0 (3.8 – 7) | 5.0 ± 0.6 (3.9 – 6.3) |
| post | 5.5 ± 0.8 (3.7 – 7.0) | 5.4 ± 0.9 (3.2 – 6.7) |

Mean ± standard deviation (min – max).

### 3.3. Characteristics of the imagery homework

The overall engagement and time investment in imagery homework were comparable between the groups (two-sample t-test, p = 0.934) with less variability across subjects in the VI group. Imagery vividness and difficulty ratings were not significantly different between the groups (two-sample t-test, p = 0.406 and 0.899, respectively). The VI group reported significantly higher rates of positive emotions (e.g., relaxed, pleasant) compared with the MI-NF group (e.g., energized, pleasant) (Mann – Whitney U Test, p < 0.001) associated with the imagery practice (Table 3).

**Table 3.** Imagery homework.

|  | **MI-NF** | **VI** |
| --- | --- | --- |
| Total time spent (min) | 320.8 ± 123.3 (110 – 540) | 323.5 ± 84.5 (192 – 528) |
| Imagery difficulty | 3.2 ± 1.3 (0.9 – 6.2) | 3.0 ± 1.3 (0.4 – 5.4) |
| Imagery vividness | 7.1 ± 1.1 (5.3 – 9.1) | 7.3 ± 1.3 (4.7 – 9.6) |
| Positive emotions (%) | 75.8 ± 12.3 (53 – 97) | 90.2 ± 11.3 (61 – 100) |

Mean ± standard deviation (min – max). Vividness and difficulty of imagery were rated on a
Likert-like scale from 0 to 10; 0: not vivid/not difficult and 10: very vivid/very difficult.

Walking, aerobic and resistance exercises, and house chores were common themes in kinesthetic motor imagery practices. Special indoor places and landscapes with mountains, lakes, etc. were common themes in visual imagery practices. Sensations of movement, heartbeat, and breathing were the most commonly reported body sensations associated with kinesthetic motor imagery. Light, color, and shape were the most commonly reported sensory modalities associated with visual imagery (see Supplementary Material Tables S5 and S6 for details).

### 3.4. Debriefing at the last visit

All subjects in the MI-NF group reported enhanced body and kinesthetic awareness as well as more presence and mindfulness during everyday movements as a result of kinesthetic motor imagery practice. One subject described this practice as “body amnesia prevention.” Increased awareness of asymmetry between the most- and least-affected sides in movement speed and range of motion was reported most frequently. This awareness prompted the subjects to make conscious corrective efforts and led to improvement in limb symmetry and coordination in movements (e.g., improved arm swing and stride length while walking, improved hook and punch while boxing, more symmetrical strokes while swimming, more coordinated upper and lower limb movements during gym exercises). About one-third of the subjects also reported improved balance while walking and putting pants or socks on. One subject reported remarkable improvement in freezing episodes while walking and jogging and was pleased by his ability to complete a road race. On the other hand, there was no noticeable improvement in fine motor skills (e.g., writing, typing, shaving, brushing teeth, playing the guitar or piano) or tremor.

### 3.5. Imaging

#### 3.5.1. Preprocessing

On average, subjects in both groups had submillimeter head motion in both resting-state and imagery scans without NF. Head motion did not differ significantly between the groups. The GCOR distribution was also not significantly different between subjects who were tested in different scanners suggesting that denoising successfully removed any potential scanner effects on the voxel-wise correlations (see Supplementary Material Tables S7 and S8).

#### 3.5.2. Task-based and resting-state functional connectivity changes

The NF sessions with short breaks in-between to reduce fatigue were overall well tolerated. Negative NF was usually associated with distraction, fatigue, thinking of the movements rather than imagining them, weak body sensations, and frustration due to imagining limitations and negative body sensations during actual movements. Positive NF was usually associated with vivid imagery with strong and positive body sensations, sustained focus, feeling in a flow state, and positive emotions (e.g., sense of accomplishment). At times, subjects felt that the NF was noncontingent with their level of effort or the same imagery practice yielded positive NF in one block and negative NF in another.

The bar graphs in Fig. 3 display the Fisher z-transformed right insula-dmFC functional connectivity strength (mean ± standard error) during rest and imagery scans with and without NF and the percent change in the right insula-dmFC functional connectivity strength (mean ± standard error) from the first to the last rest and imagery scans without NF for both groups. There was significant between-subject variability in all functional connectivity measures in both groups. Between-group comparisons of the percent change in right insula-dmFC functional connectivity strength from the first to the last rest and imagery scans without NF did not reveal a significant difference (Mann-Whitney U test, p = 0.481 and 0.981, respectively).

**Fig. 3.**
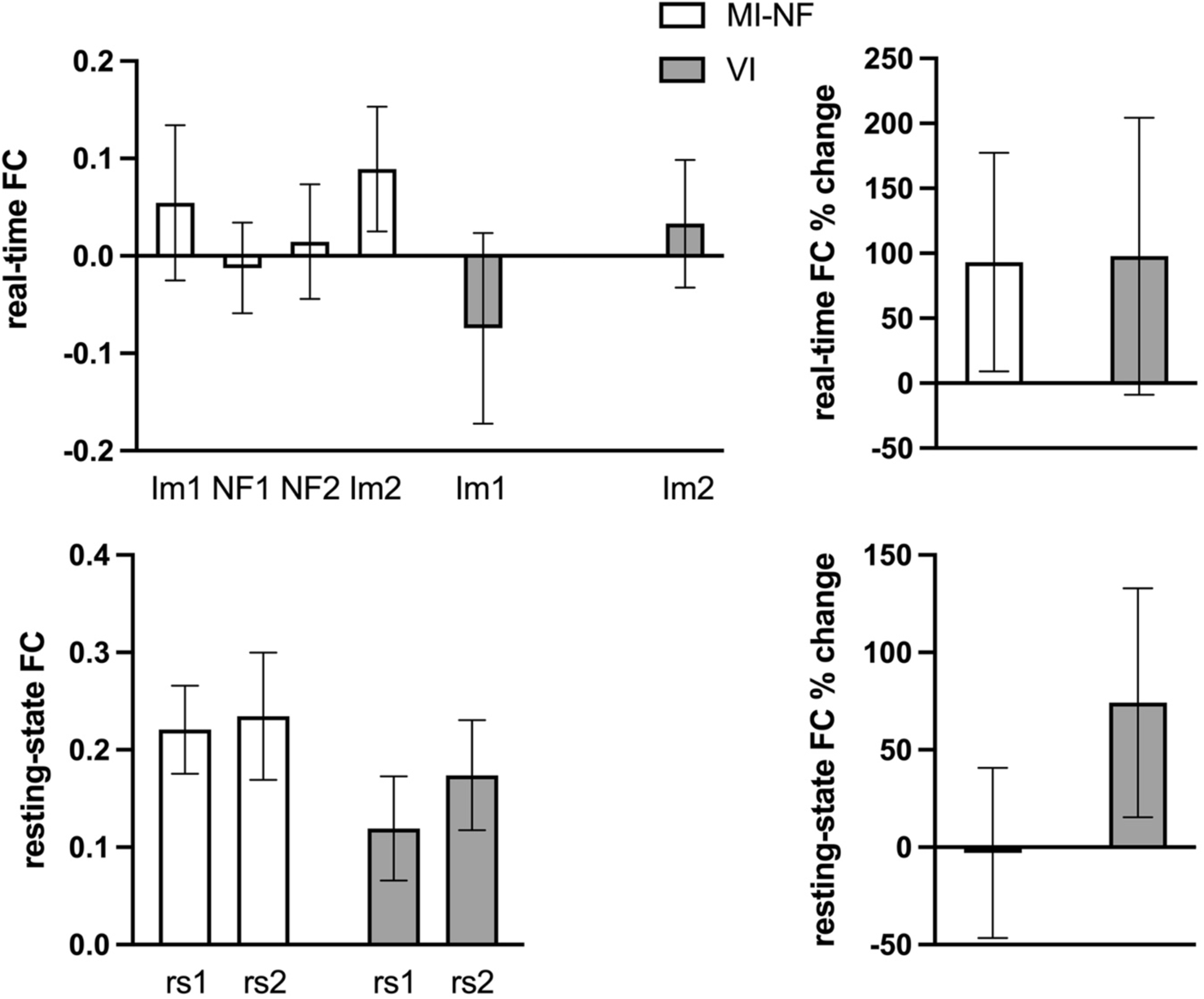
Right insula-dmFC functional connectivity. Error bars show the standard error. FC: Functional connectivity, Im1 and Im2: first and last respective imagery scans without neurofeedback, NF1 and NF2: Neurofeedback scans on days 1 (visit 2) and 2 (visit 3), rs1 and rs2: first and last resting-state scans. *MI-NF*: Im1: 0.05 ± 0.08, NF1: −0.01 ± 0.05, NF2: 0.01 ± 0.06, Im2: 0.09 ± 0.06, real-time % change: 93.2 ± 84.1, rs1: 0.22 ± 0.05, rs2: 0.23 ± 0.07, resting-state % change: −2.9 ± 43.7. *VI*: Im1: −0.07 ± 1.00, Im2: 0.03 ± 0.07, real-time % change: 97.8 ± 106.6, rs1: 0.12 ± 0.05, rs2: 0.17 ± 0.06, resting-state % change: 74.2 ± 58.8.

#### 3.5.3. Correlations between functional connectivity and motor performance changes

The Kendall’s tau-b correlation between the average percent change in motor performance scores and percent change in imagery task-based right insula-dmFC functional connectivity was significant in the MI-NF group (correlation coefficient = 0.307, p = 0.045, 95% CI [0.012, 0.553]) (Fig. S1) and not significant in the VI group (correlation coefficient = −0.108, p = 0.481, 95% CI [−0.392, 0.194]).

The Kendall’s tau-b correlation between the average percent change in motor performance scores and percent change in resting-state right insula-dmFC functional connectivity was not significant in the MI-NF (correlation coefficient = 0.273, p = 0.076, 95% CI [−0.26, 0.526]) or the VI (correlation coefficient = −0.195, p = 0.204, 95% CI [−0.464, 0.018]) group.

The Kendall’s tau-b correlation between the average percent change in MDS-UPDRS part III scores and percent change in imagery task-based right insula-dmFC functional connectivity was not significant in the MI-NF group (correlation coefficient = −0.035, p = 0.821, 95% CI [−0.328, 0.264]) or the VI group (correlation coefficient = −0.091, p = 0.554, 95% CI [−0.377, 0.211]).

The Kendall’s tau-b correlation between the average percent change in MDS-UPDRS part III scores and percent change in resting-state right insula-dmFC functional connectivity was not significant in the MI-NF (correlation coefficient = 0.017, p = 0.910, 95% CI [−0.280, 0.312]) or the VI (correlation coefficient = −0.091, p = 0.554, 95% CI [−0.377, 0.211]) group.

#### 3.5.4. Whole-brain resting-state functional connectivity

The VI group did not show any significant pairwise whole-brain functional connectivity differences in the rest 2 > rest 1 contrast. The MI-NF group showed significantly reduced pairwise whole-brain functional connectivity in the rest 2 > rest1 contrast between several ventral and dorsal frontal nodes, and between the temporal pole and caudate nodes, and increased connectivity between a few cerebellar and fronto-temporal nodes. There was a significant group × time interaction (MI-NF > VI and rest 2 > rest 1 contrast) demonstrating decreased functional connectivity between only a few cerebellar nodes and increased functional connectivity between a cerebellar node and orbitofrontal nodes (see Supplementary Material Fig. S2 and Table S9).

#### 3.5.5. Whole-brain imagery task-based functional connectivity

Both groups demonstrated significant pairwise functional connectivity changes as a result of training (imagery 2 > imagery 1 contrast) predominantly in the form of decreased functional connectivity. The main effects of group are summarized in the Supplementary Material Fig. S3 and S4 and Tables S10 and S11. Here, we focus on the interaction effects, namely the MI-NF > VI and imagery 2 > imagery 1 contrast (Fig. 4 and Table 4). As a result of kinesthetic motor imagery training, the MI-NF group showed stronger functional connectivity in cerebellar, occipito-temporal, premotor, and frontal nodes, as well as the insula and midbrain. As a result of visual imagery training, the VI group showed stronger functional connectivity in a different set of cerebellar, occipito-temporal, and frontal nodes, as well as posterior cingulate nodes and pons.

**Fig. 4.**
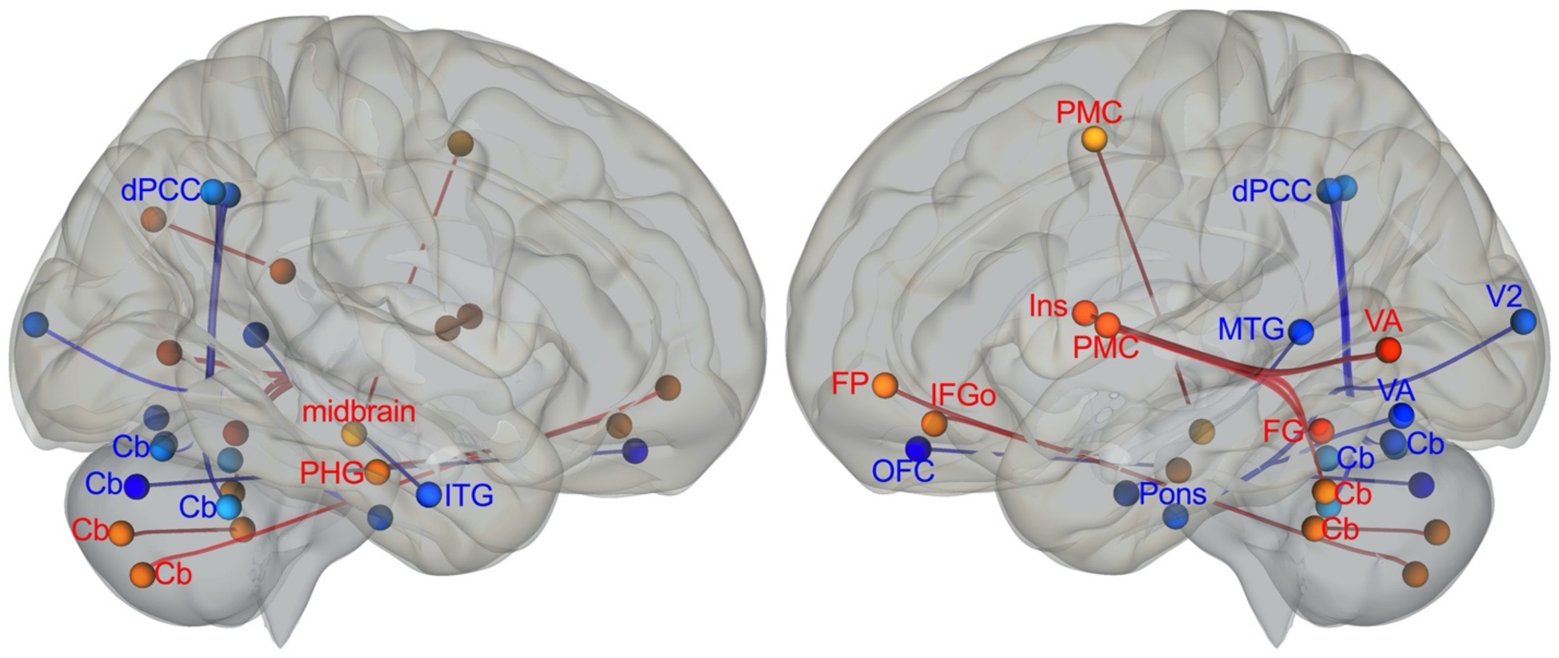
Whole-brain imagery task-based functional connectivity changes. Red: MI-NF > VI and blue: VI > MI-NF. Cb: Cerebellum, dPCC: Dorsal posterior cingulate cortex, FG: Fusiform gyrus, FP: Frontal pole, IFGo: Inferior frontal gyrus, orbital part; Ins: Insula, ITG: Inferior temporal gyrus, MTG: Middle temporal gyrus, OFC: Orbitofrontal cortex, PHG: Parahippocampal gyrus, PMC: Premotor cortex, V2: Secondary visual cortex, VA: Visual association cortex.

**Table 4.**
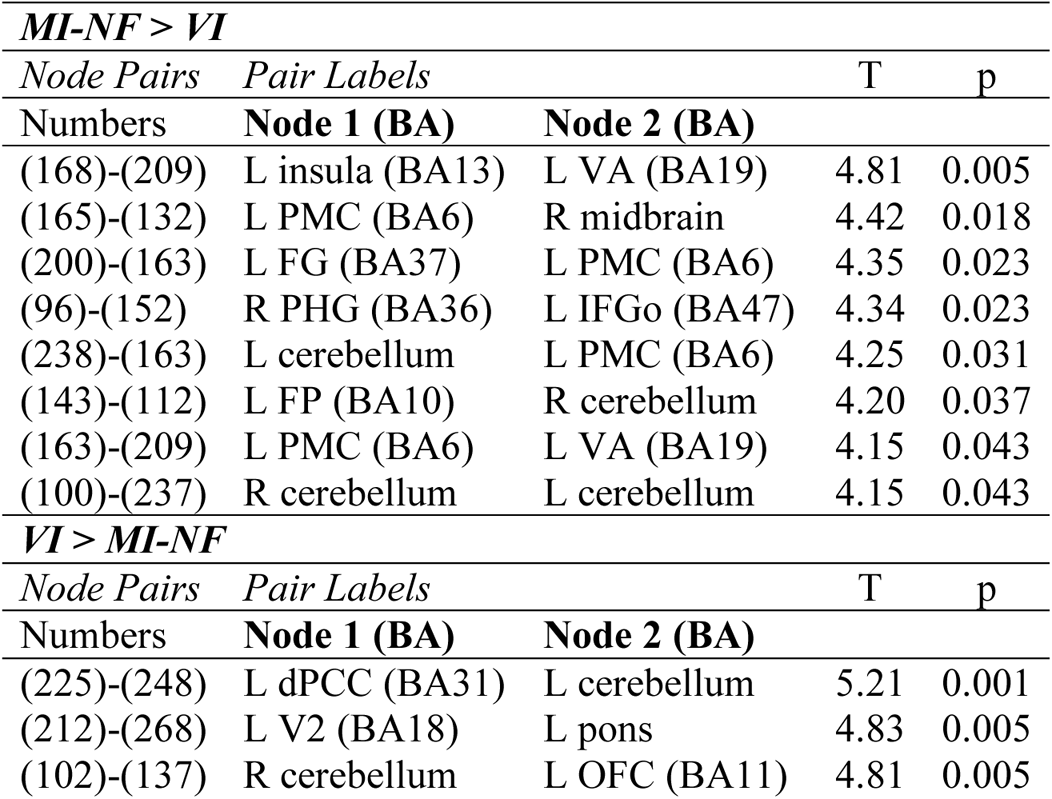
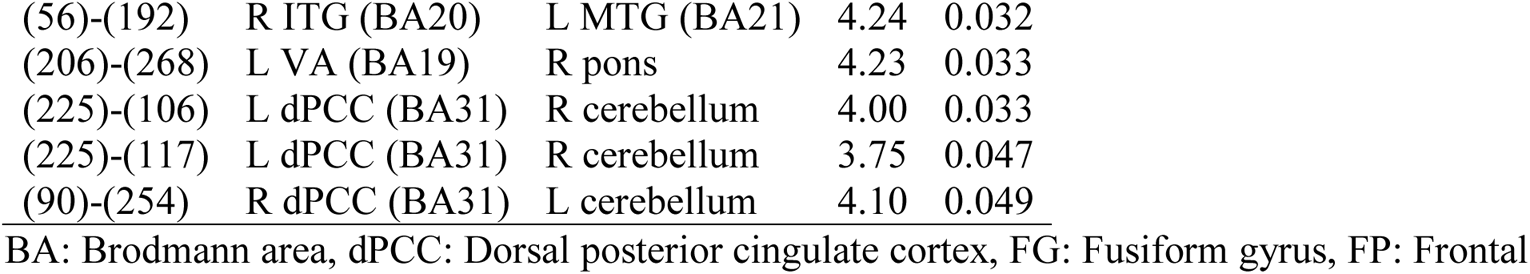

## 4. Discussion

In summary, the primary imaging and clinical outcomes of our study did not reach statistical significance. However, we did see significant improvements in the secondary clinical outcome where both groups receiving motor imagery (MI-NF) and visual imagery (VI) improved their composite gross motor function scores comparably. Notably, there was a significant positive relationship between the change in real-time right insula-dmFC functional connectivity and improvement in gross motor function scores only in the MI-NF group. There was no significant relationship between other imaging and clinical outcomes. The exploratory outcome measures showed statistical significance. Both groups reported significant subjective improvement in their imagery skills as hypothesized. The whole-brain task-based functional connectivity patterns mapped onto distinct networks reflecting the specific type of imagery training (i.e., kinesthetic motor vs visual) in each group.

### 4.1. Imaging outcomes: Heterogeneity in NF regulation success

Contrary to our findings based on eight subjects in the pilot study (Tinaz et al., 2018), we did not find significant NF regulation in the MI-NF group. A high degree of variability in NF response in healthy controls and clinical populations is not uncommon. It is estimated that about 30% of participants fail to learn how to regulate their brain signal with NF (Sorger et al., 2019), and studies report mixed results regarding overall regulation success (about 30% of studies) and behavior change (about 50% of studies) (Thibault et al., 2018). There are numerous methodological and subjective factors contributing to this variability (Sorger et al., 2019). During post-scan debriefing, we identified nonspecific factors including mental fatigue and distraction, and subject-specific factors such as frustrated efforts during imagery as potential contributors to NF regulation failure. This is not surprising given that the insula is an essential hub in the appraisal of emotional valence (Craig 2009), and negative emotions during motor imagery would be expected to interfere with the regulation of insula activity and co-activity with the dmFC.

Another potential factor was the choice of mental strategy during NF training. We instructed the subjects to use an explicit kinesthetic motor imagery strategy emulating everyday complex motor activities with the aim of maximizing clinical benefit. However, using spontaneous mental strategies as opposed to following explicit instructions can be more efficient to achieve successful NF modulation (Kober et al., 2013). For example, NF training with explicit motor imagery instructions has been shown to be ineffective in up-regulation of the SMA activity in a small group of healthy controls (Sepulvada et al., 2016). Furthermore, in a graded NF paradigm combined with kinesthetic hand motor imagery, subjects could up-regulate the SMA activity to the specified level, but no interaction between target level and feedback condition was found suggesting a general motor imagery effect, but no specific feedback modulation in the SMA (Mehler et al., 2019).

Finally, the number of NF training runs (a total of 10-12 per subject) may not have been sufficient for all subjects to ensure successful regulation.

### 4.2. Clinical outcomes: Scale sensitivity and training-specific vs nonspecific effects

There was no clinically important difference in the MDS-UPDRS part III scores in the MI-NF group post-training. The total MDS-UPDRS part III score is the gold standard primary outcome measure in clinical trials in PD. Tremor and rigidity (muscle stiffness) are major components of the scale, which were not specific targets of the motor imagery training. Bradykinesia (slowness) is another major component, also a training target, but is rated using rather simple repetitive movements. Therefore, part III may not have been sensitive enough to the changes in complex movements. These changes were captured by the motor function tests that we used to measure the speed of complex movements and activities of daily living. Both groups improved their motor function scores significantly and comparably suggesting a nonspecific or placebo effect as a result of participating in an interventional study. This is particularly relevant for individuals with PD, who also demonstrate strong placebo effects in clinical trials with pharmacological and surgical interventions (Lidstone 2014). Our study design did not allow us to exclude the placebo effect, however, we think that the motor improvement in the MI-NF group may also reflect some specificity. The modestly significant positive correlation between the change in real-time right insula-dmFC functional connectivity and improvement in motor function scores was observed only in the MI-NF group suggesting training-specific effects. An alternative explanation for the motor function improvement in the VI group would be “spillover” effects. The visual imagery practice required sustained attention, multisensory integration, and visuospatial construction (Pearson 2019). Moreover, subjects in the VI group reported significantly higher rates of positive emotions (predominantly “relaxed”) associated with their daily practice compared with the MI-NF group. Taken together, the visual imagery practice may have indirectly enhanced motor functioning by promoting relaxed alertness and sustained visuospatial attention.

### 4.3. Exploratory outcomes: Training effects on behavior and brain connectivity

Though the MIQ-3 is a movement imagery questionnaire, the imagery task includes two visualization conditions from a first- and third-person perspective, and a kinesthetic condition. Therefore, it is not surprising that both groups reported significant improvement in the MIQ-3 post-training. The MI-NF group also reported enhanced kinesthetic awareness and mindfulness during everyday movements as a result of practice, which led to corrective efforts and improvement in movement size, speed, coordination, and balance. These were subjective accounts gathered at the post-training debriefing, therefore, could only be analyzed qualitatively. Many NF studies with neuropsychiatric populations use self-evaluation surveys as outcome measures of symptomatic improvement underlining the need to develop specific surveys to assess changes in kinesthetic awareness of complex movements quantitatively.

Notably, we did see specificity by the MI-NF versus the VI training (imagery training effects: imagery 2 > imagery 1 contrast) mapped onto separate whole-brain functional connectivity patterns during the respective imagery scans without NF. Of note, the whole-brain intrinsic functional connectivity changes (rest 2 > rest 1 contrast) were not robust and did not express a clear pattern suggesting that the training effects were specific to task performance. Post-training, the MI-NF group compared with the VI group exhibited stronger functional connectivity between the nodes involved in kinesthetic motor imagery including the insula and premotor cortex, whereas the VI group compared with the MI-NF group demonstrated stronger functional connectivity between the nodes involved in visual imagery including nodes in the ventral and dorsal visual streams. These results provide a manipulation check and validate that the training targeted the expected brain networks, and are also in line with previous reports of differential activation patterns during kinesthetic motor (Guillot et al., 2009; Hétu et al., 2013) and visual imagery (Pearson 2019) corroborating that subjects in both groups adhered to the specific imagery instructions in their practice.

Different parts of the cerebellum show intrinsic functional connectivity with different large-scale brain networks and are involved in a range of sensorimotor and cognitive tasks (Buckner et al., 2011). Different cerebellar nodes were also recruited in both imagery tasks demonstrating increased functional connectivity with the premotor and frontal nodes in the motor imagery task, and with the orbitofrontal (OFC) and dorsal posterior cingulate cortex (dPCC) in the visual imagery task. The OFC is considered a hub for integrating multimodal sensory information with hedonic value (Kringelbach 2005) and may have been involved in the retrieval of multimodal sensory information and integrating it with subjects’ emotional experience during visual imagery. The dPCC has rich anatomical connections with temporal, parietal, frontal regions and is thought to regulate attention according to cognitive demands (Leech and Sharp, 2014). During visual imagery, the dPCC may have been involved in monitoring the retrieval of episodic memories and visuospatial construction of scenes.

In addition to the insula and premotor nodes, increased functional connectivity in the motor imagery network also included the fusiform and parahippocampal nodes, which may have played a role in imagining one’s body movements (Olsson et al., 2008) in specific environmental contexts (Epstein et al., 2017), respectively. Finally, a brainstem node overlapping with the midbrain/red nucleus region also displayed increased functional connectivity with the premotor cortex during motor imagery. The midbrain/red nucleus is an important subcortical region in motor control especially of the limbs, motor coordination, and motor preparation through its connections with the motor cortices, basal ganglia, and cerebellum (Basile et al., 2021).

Taken together, our results demonstrate the neural bases of task-specific practice effects over the course of four weeks in the form of functional re-organization of distinct neural circuits supporting kinesthetic motor and visual imagery. However, we cannot rule out the potential large-scale neuromodulatory effects of NF training in the MI-NF group. At least indirectly, subjects’ experience during NF sessions and post-scan debriefing informed their practice strategies at home and may have helped them hone their kinesthetic motor imagery skills with practice.

## Limitations

Involuntary muscle contraction as a result of motor imagery can potentially confound the BOLD signal (Thibault et al., 2018). We did not monitor muscle activity in the scanner using surface EMG electrodes. Though we did not observe overt movements while subjects were in the scanner and head motion levels in the scanner across both groups were equivalent, we could not rule out covert muscle contractions. We also did not track eye movements during imagery blocks, thus, could not assess whether subjects in both groups were matched in the amount of eye movement. The training and assessment of outcome measures were limited to a four-week period with no follow-up. Continued symptomatic benefits for weeks after NF have been demonstrated in clinical populations (Rance et al., 2018).

## Conclusions

The functional connectivity-based NF regulation was unsuccessful in our cohort with mild PD. The heterogeneity in NF response in healthy controls and particularly in clinical populations is an ongoing challenge in the field, and concerted efforts are made to address the potential sources of heterogeneity (e.g., selecting the appropriate NF targets, dose, mental strategies, study designs, and meaningful outcome measures, and ways to predetermine potential success). Yet, kinesthetic motor imagery practice by itself is a promising intervention in PD. There is also evidence that multisensorial integrative imagery techniques (e.g., dynamic-cognitive imagery) (Abraham et al., 2018) and motor imagery combined with action observation (Caligiore et al., 2017) may improve motor functions in people with PD. Future studies are needed to develop specific motor imagery protocols and determine practice parameters (e.g., dose, duration, outcome measures), which then can be individualized and incorporated into clinical practice as compensatory rehabilitation strategies for motor impairment in people with PD.

## CRediT authorship contribution statement

**Sule Tinaz**: Conceptualization, Methodology, Investigation, Formal Analysis, Visualization, Supervision, Writing – original draft, Funding Acquisition. **Serageldin Kamel**: Investigation, Formal Analysis, Data Curation, Visualization. **Sai S. Aravala**: Investigation, Formal Analysis.. **Mohamed Elfil**: Investigation, Formal Analysis. **Ahmed Bayoumi**: Investigation, Formal Analysis. **Amar Patel**: Investigation, Formal Analysis. **Dustin Scheinost**: Methodology, Software. **Rajita Sinha**: Conceptualization, Methodology, Supervision. **Michelle Hampson**: Conceptualization, Methodology, Software, Resources. All authors were involved in the review and editing of the manuscript.

## Data Availability

All data produced in the present study are available upon reasonable request to the authors

## Acknowledgment

This work was supported by the National Center for Advancing Translational Science, a component of the National Institutes of Health (grant numbers UL1TR001863, KL2TR001862) and by the National Institute of Neurological Disorders and Stroke (grant number K23NS099478). We thank Jessica Lemere and Sayan Basu for their help with data collection, and Jessica Lemere for her help with imagery audio recordings.

## Supplementary Material

### 1. Eligibility

Inclusion Criteria:

- Subjects with a diagnosis of idiopathic PD defined according to the UK Brain Bank diagnostic criteria
- Age ≥ 40 years
- Expected to be on a stable dopaminergic medication regimen throughout the study period

Exclusion Criteria:

- Age < 40 years
- Non-English speaking
- Pregnancy
- Breastfeeding
- Excessive alcohol consumption (> 7 drinks per week for women, > 14 drinks per week for men) or illicit substance use
- History of a neurological disorder such as a brain tumor, stroke, central nervous system infection, multiple sclerosis, movement disorder (other than PD), or seizures
- History of schizophrenia, bipolar disorder, attention deficit disorder, or obsessive-compulsive disorder
- History of head injury with loss of consciousness longer than a minute
- Metallic surgical implants or traumatically implanted metallic foreign bodies
- Inability to lie flat for about an hour in the MRI scanner
- Discomfort being in small, enclosed spaces
- Dementia (Montreal Cognitive Assessment score < 21)
- Depression (Beck Depression Inventory-II score > 28 indicating more than moderate depression)
- Hoehn & Yahr stage > 3
- Focal neurological findings on exam that suggest cerebral pathology other than that associated with parkinsonism
- Motor symptoms that could potentially introduce too much motion artifact in the imaging data (e.g., Movement Disorders Society-Unified Parkinson’s Disease Rating Scale resting tremor score > 2 in limbs, head/chin tremor, or more than mild dyskinesia by history or exam).
- Poor homework compliance (< 50% completion rate).

### 2. Clinical and psychometric results

**Table S1.** Normative mean and cutoff scores for each test.

|  |  |
| --- | --- |
| Apathy | $> 14$ significant apathy |
| MoCA – dementia | $< 21$ |
| MoCA – mild cognitive impairment | < 26 |
| BDI-II minimal | 0 – 13 |
| BDI-II mild | 14 – 19 |
| BDI-II moderate | 20 – 28 |
| STAI-S male | >34.51 significant state anxiety |
| STAI-S female | >32.20 significant state anxiety |
| STAI-T male | >33.86 significant trait anxiety |
| STAI-T female | >31.79 significant trait anxiety |
| PFS-16 | >2.95 significant fatigue |
| PDQ39-SI | >31.6 significantly poor quality of life summary index for those with disease stage of H&Y 2 |

#### 2.1. Comparisons with normative data

**MI-NF group**: The distribution of the MoCA and apathy scores was not normal. We used the median scores of MoCA and apathy scale, and mean scores of the rest for comparison with normative data:

MoCA: The median score (29) was significantly above the MCI cutoff (p = 0.000).

STAI-S: Males (mean: 30.33) were not significantly different from (p = 0.058) and females (mean: 28.10) showed significantly less state anxiety (p = 0.014) than the age-matched normal population.

STAI-T: Males (mean: 33.83) and females (mean: 32.70) were not significantly different from the age-matched normal population in trait anxiety (p = 0.989 and 0.717, respectively).

BDI-II: The mean score was significantly below the mild depression cutoff (p = 0.000). Apathy: The median score (5) was significantly below the apathy cutoff (p = 0.000).

PFS-16: The mean score was significantly below the fatigue cutoff (p = 0.002).

PDQ39-SI: The mean score was significantly below the cutoff for poor quality of life (p = 0.000).

**VI group**: The distribution of the MoCA, STAI-S, STAI-T, BDI-II, PFS-16, and PDQ39-SI was not normal. We used the median scores for comparison with normative data:

MoCA: The median score (29) was significantly above the MCI cutoff (p = 0.000).

STAI-S: Males (median: 31.50) were not significantly different (p = 0.192) and females (median: 26.00) showed significantly less state anxiety (p = 0.006) than the age-matched normal population.

STAI-T: Males (median: 34.00) and females (median: 30.50) were not significantly different from the age-matched normal population in trait anxiety (p = 0.953 and 0.389, respectively). BDI-II: The median score (6) was significantly below the mild depression cutoff (p = 0.000). Apathy: The mean score was significantly below the apathy cutoff (p = 0.000).

PFS-16: The median score (2.095) was significantly below the fatigue cutoff (p=0.000).

PDQ-39 SI: The median score (12.32) was significantly below the cutoff for poor quality of life (p=0.000).

#### 2.2. Between-group comparisons

The two-sample t-test on normally distributed variables, Mann-Whitney U test on non-normally distributed variables, and Chi-Square test for handedness and disease onset side did not reveal any significant difference between the MI-NF and VI groups.

**Table S2.** Between-group statistics p values.

|  | **Mann-Whitney U** | **Two-sample t** | **Chi-square** |
| --- | --- | --- | --- |
| Age |  | 0.861 |  |
| Handedness |  |  | 0.380 |
| Onset side |  |  | 0.763 |
| H & Y |  | 0.561 |  |
| Disease duration | 0.869 |  |  |
| MDS-UPDRS I |  | 0.459 |  |
| MDS-UPDRS II |  | 0.152 |  |
| MDS-UPDRS III |  | 0.400 |  |
| MDS-UPDRS IV | 0.267 |  |  |
| MoCA | 0.689 |  |  |
| STAI-S | 0.944 |  |  |
| STAI-T | 0.897 |  |  |
| BDI-II | 0.613 |  |  |
| Apathy | 0.207 |  |  |
| PFS-16 | 0.814 |  |  |
| PDQ39-SI | 0.916 |  |  |

**Table S3.** Medications.

|  | **Number of subjects** |  |
| --- | --- | --- |
|  | **MI-NF** | **VI** |
| **Type of medication** |  |  |
| Carbidopa/levodopa | 14 | 16 |
| Dopamine receptor agonists | 4 | 6 |
| Monoamine oxidase-B inhibitors | 13 | 10 |
| Amantadine | 5 | 5 |
| Mucuna powder | 1 | - |
| **Combination of medications** |  |  |
| 0 (no medication) | 2 | 5 |
| 1 medication | 6 | 6 |
| 2 medications | 11 | 4 |
| 3 medications | 3 | 6 |
| 4 medications | - | - |
| 5 medications | - | 1 |

**Table S4.** LEDD distribution.

| **LEDD (mg)** | **MI-NF** | **VI** |
| --- | --- | --- |
| Visit 1 | 323.1 ± 242.6 (0.0 – 870.0) | 439.9 ± 395.6 (0.0 – 1640.0) |
| Visit 2 | 336.0 ± 243.4 (0.0 – 870.0) | 414.4 ± 405.6 (0.0 – 1640.0) |
| Visit 3 | 329.8 ± 221.6 (0.0 – 700.0) | 417.2 ± 400.9 (0.0 – 1640.0) |
| Visit 4 | 332.6 ± 218.4 (0.0 – 700.0) | 385.9 ± 367.6 (0.0 – 1640.0) |

Mean ± standard deviation (min – max). Repeated measures ANOVA (within-subject: visits with four levels, between-subject: group) did not reveal any significant effect of visit ( $F(1,40) = 0.359$ , $p = 0.783$ ), group ( $F(1,42) = 0.776$ , $p = 0.383$ ), or visit x group interaction ( $F(1,40) = 1.247$ , $p = 0.305$ ).

**Table S5.** MI-NF group imagery homework.

| Activity | % | Sensation | % | Emotion | % |
| --- | --- | --- | --- | --- | --- |
| Walking | 18.4 ± 11.2 | Movement | 20.0 ± 7.1 | *Positive* |  |
| Calisthenics | 13.1 ± 11.0 | Heart beat | 15.3 ± 6.5 | Energized | 20.2 ± 10.9 |
| Weights | 9.7 ± 11.2 | Breathing | 14.6 ± 4.4 | Pleasant | 18.1 ± 6.3 |
| Indoor ADL | 9.2 ± 6.7 | Stretch | 10.2 ± 4.9 | Comfortable | 16.1 ± 7.8 |
| Outdoor ADL | 8.3 ± 9.1 | Warmth/coolness | 9.2 ± 5.7 | Refreshed | 8.3 ± 4.1 |
| Balancing | 8.2 ± 9.0 | Tightness/looseness | 8.2 ± 4.0 | Relaxed | 7.0 ± 4.0 |
| Boxing | 7.8 ± 12.1 | Heaviness/lightness | 5.5 ± 3.2 | Exhilarated | 5.9 ± 3.2 |
| Swimming | 7.0 ± 7.5 | Touch | 4.4 ± 3.6 | *Negative* |  |
| Running | 3.9 ± 5.9 | Pressure | 4.2 ± 3.4 | Tired | 14.2 ± 9.1 |
| LSVT | 3.7 ± 9.6 | Stiffness | 3.6 ± 2.8 | Uncomfortable | 3.3 ± 3.1 |
| Treadmill | 3.0 ± 5.1 | Pain | 2.4 ± 2.0 | Frustrated | 2.4 ± 2.9 |
| Golfing | 1.7 ± 5.1 | Burning | 1.1 ± 1.5 | Unpleasant | 1.9 ± 2.2 |
| Skiing | 1.4 ± 4.6 | Tremor | 1.0 ± 2.0 | Bothered | 1.2 ± 2.0 |
| Dressing | 1.0 ± 2.6 | Dizziness | 0.3 ± 0.8 | Distracted | 1.2 ± 1.6 |
|  |  | Tingling | 0.3 ± 0.8 |  |  |

ADL: Activities of daily living, indoors: e.g., cleaning, washing dishes, doing laundry; outdoors: e.g., gardening, raking leaves, shoveling snow. Mean% ± standard deviation.

**Table S6.**
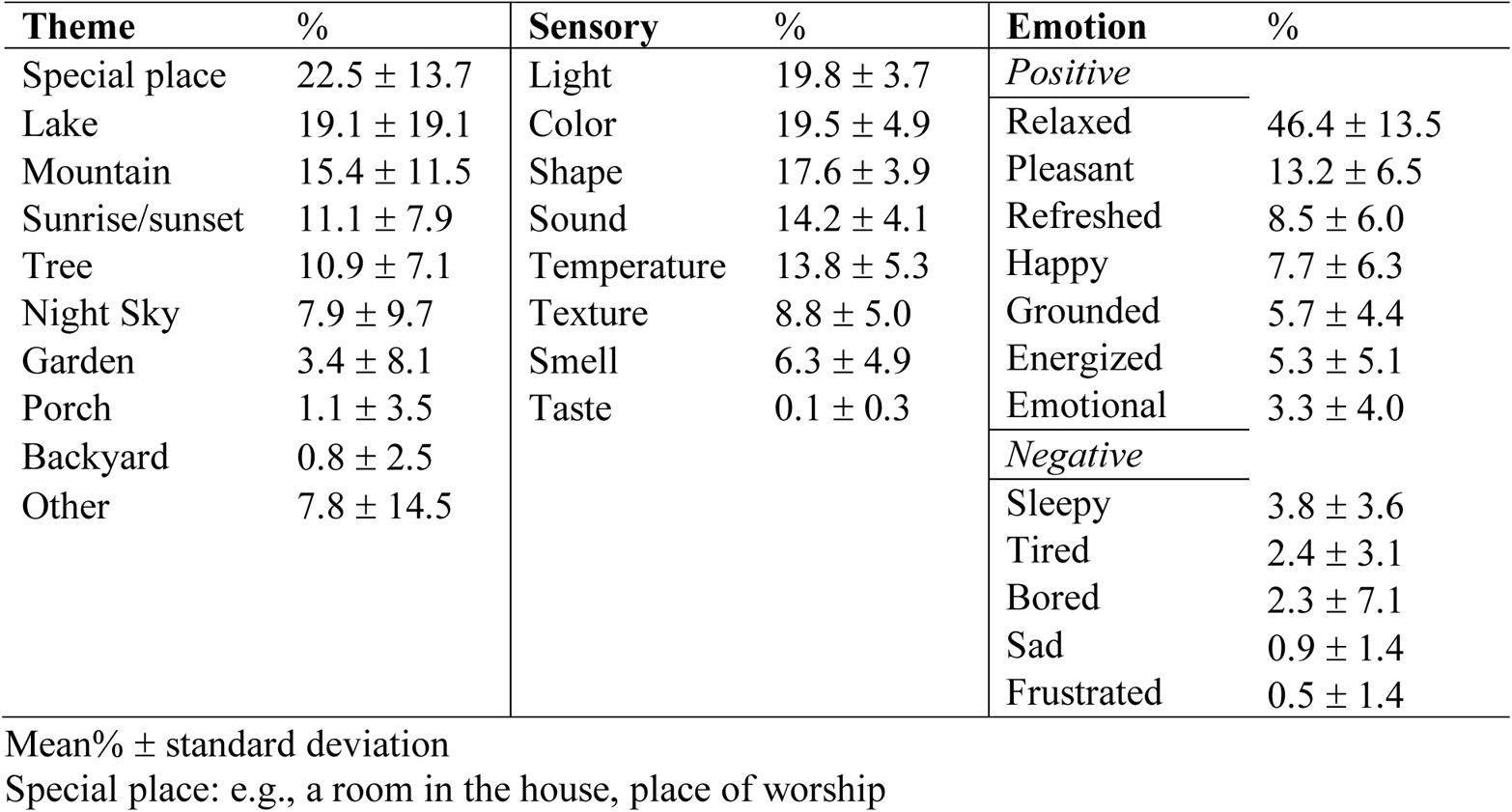
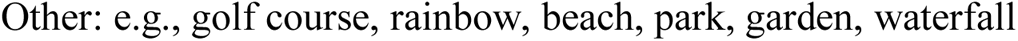
VI group imagery homework.

| Theme | % | Sensory | % | Emotion | % |
| --- | --- | --- | --- | --- | --- |
| Special place | 22.5 ± 13.7 | Light | 19.8 ± 3.7 | *Positive* |  |
| Lake | 19.1 ± 19.1 | Color | 19.5 ± 4.9 | Relaxed | 46.4 ± 13.5 |
| Mountain | 15.4 ± 11.5 | Shape | 17.6 ± 3.9 | Pleasant | 13.2 ± 6.5 |
| Sunrise/sunset | 11.1 ± 7.9 | Sound | 14.2 ± 4.1 | Refreshed | 8.5 ± 6.0 |
| Tree | 10.9 ± 7.1 | Temperature | 13.8 ± 5.3 | Happy | 7.7 ± 6.3 |
| Night Sky | 7.9 ± 9.7 | Texture | 8.8 ± 5.0 | Grounded | 5.7 ± 4.4 |
| Garden | 3.4 ± 8.1 | Smell | 6.3 ± 4.9 | Energized | 5.3 ± 5.1 |
| Porch | 1.1 ± 3.5 | Taste | 0.1 ± 0.3 | Emotional | 3.3 ± 4.0 |
| Backyard | 0.8 ± 2.5 |  |  | *Negative* |  |
| Other | 7.8 ± 14.5 |  |  | Sleepy | 3.8 ± 3.6 |
|  |  |  |  | Tired | 2.4 ± 3.1 |
|  |  |  |  | Bored | 2.3 ± 7.1 |
|  |  |  |  | Sad | 0.9 ± 1.4 |
|  |  |  |  | Frustrated | 0.5 ± 1.4 |

Mean% ± standard deviation
Special place: e.g., a room in the house, place of worship
Other: e.g., golf course, rainbow, beach, park, garden, waterfall

**Table S7.** Head Motion.

|  | MI-NF | VI | Mann-Whitney<br>U test *p* values |
| --- | --- | --- | --- |
| **Resting-state scans** |  |  |  |
| # Invalid scans | 3.23 ± 6.02 | 4.27 ± 9.21 | 0.755 |
| Maximum translation (mm) | 0.77 ± 0.50 | 0.93 ± 0.86 | 0.589 |
| Mean translation (mm) | 0.17 ± 0.07 | 0.18 ± 0.07 | 0.526 |
| **Imagery scans without NF** |  |  |  |
| # Invalid scans | 1.64 ± 2.13 | 1.5 ± 2.61 | 0.570 |
| Maximum translation (mm) | 0.74 ± 0.60 | 0.79 ± 0.90 | 0.833 |
| Mean translation (mm) | 0.16 ± 0.06 | 0.16 ± 0.07 | 0.851 |

Mean ± standard deviation. The motion variables were not normally distributed. Therefore, nonparametric Mann-Whitney U test was used for between-group comparisons, which did not reveal a significant difference between the groups.

**Table S8.**
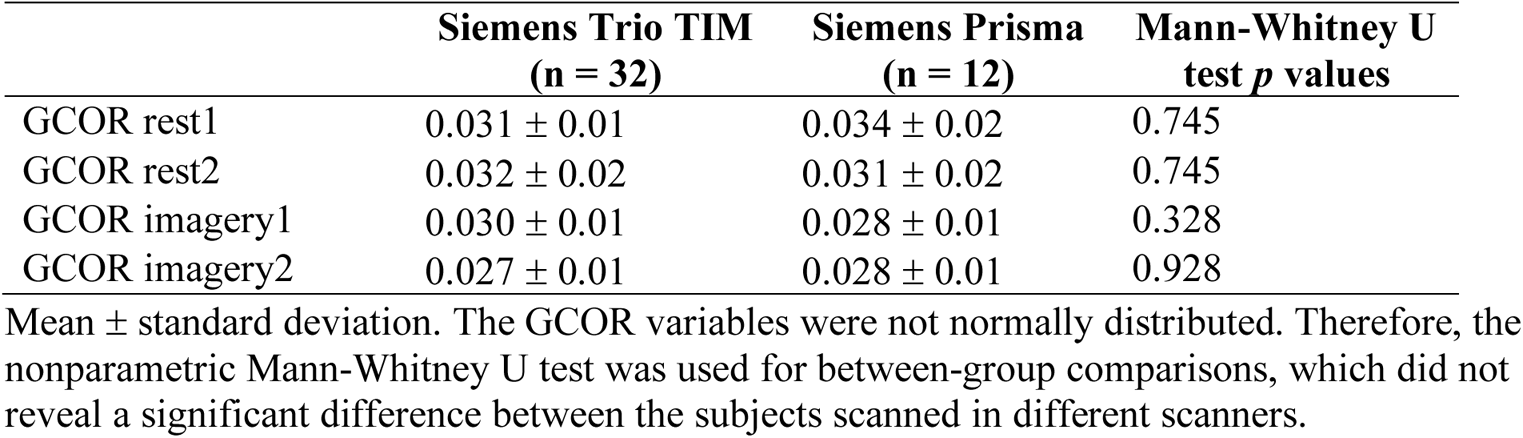
Voxel-wise global mean correlations (GCOR) per scanner after denoising.

|  | Siemens Trio TIM<br>(n = 32) | Siemens Prisma<br>(n = 12) | Mann-Whitney U<br>test *p* values |
| --- | --- | --- | --- |
| GCOR rest1 | 0.031 ± 0.01 | 0.034 ± 0.02 | 0.745 |
| GCOR rest2 | 0.032 ± 0.02 | 0.031 ± 0.02 | 0.745 |
| GCOR imagery1 | 0.030 ± 0.01 | 0.028 ± 0.01 | 0.328 |
| GCOR imagery2 | 0.027 ± 0.01 | 0.028 ± 0.01 | 0.928 |

Mean ± standard deviation. The GCOR variables were not normally distributed. Therefore, the nonparametric Mann-Whitney U test was used for between-group comparisons, which did not reveal a significant difference between the subjects scanned in different scanners.

**Fig. S1.**
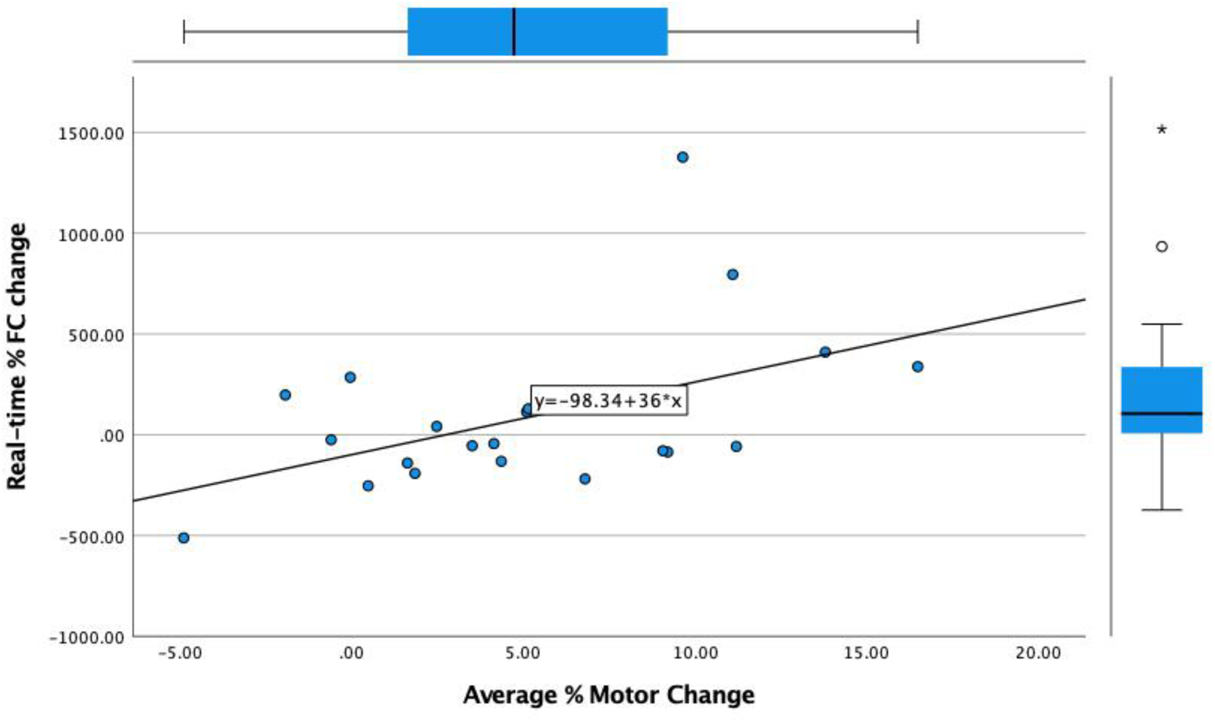
Relationship between the imaging and motor outcomes in the MI-NF group. The graph displays the relationship between the average percent change in motor performance scores and percent change in real-time imagery task-based right insula-dmFC functional connectivity in the MI-NF group.

**Fig. S2.**
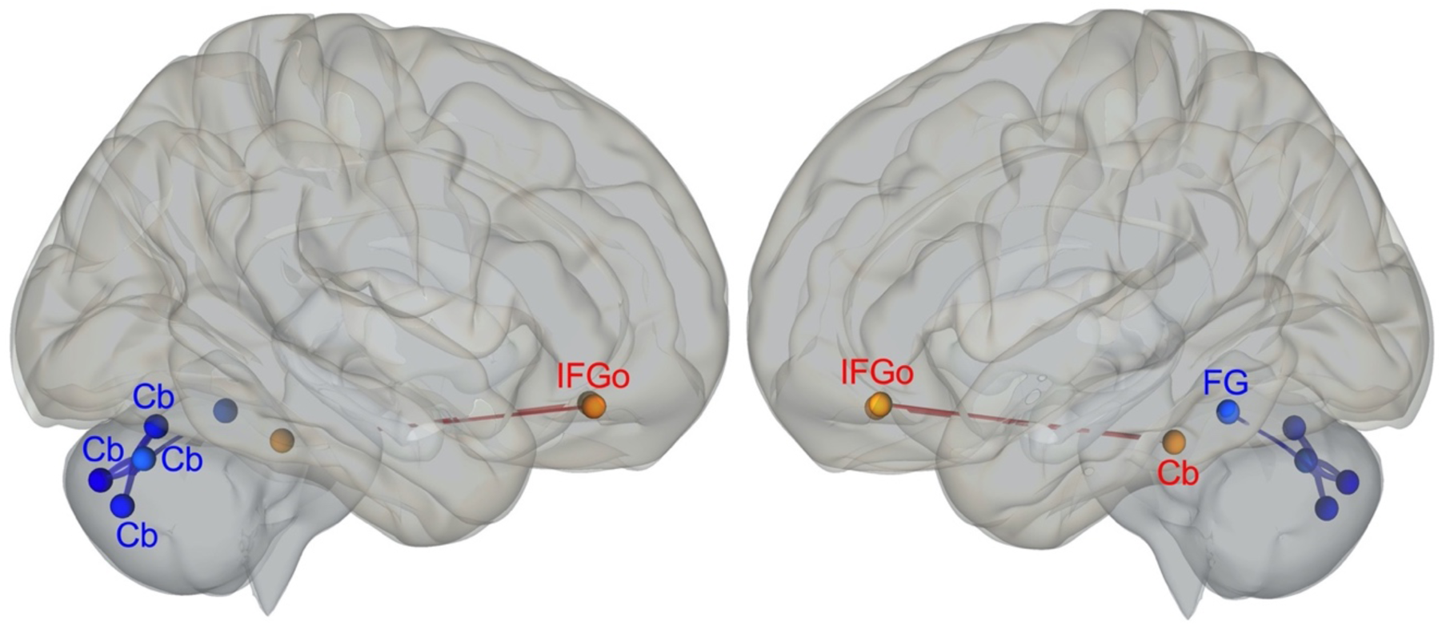
Whole-brain resting-state functional connectivity: MI-NF > VI and rest2 > rest1. Red: increased, blue: decreased. Cb: Cerebellum, FG: Fusiform gyrus, IFGo: Inferior frontal gyrus, orbital part.

**Table S9.**
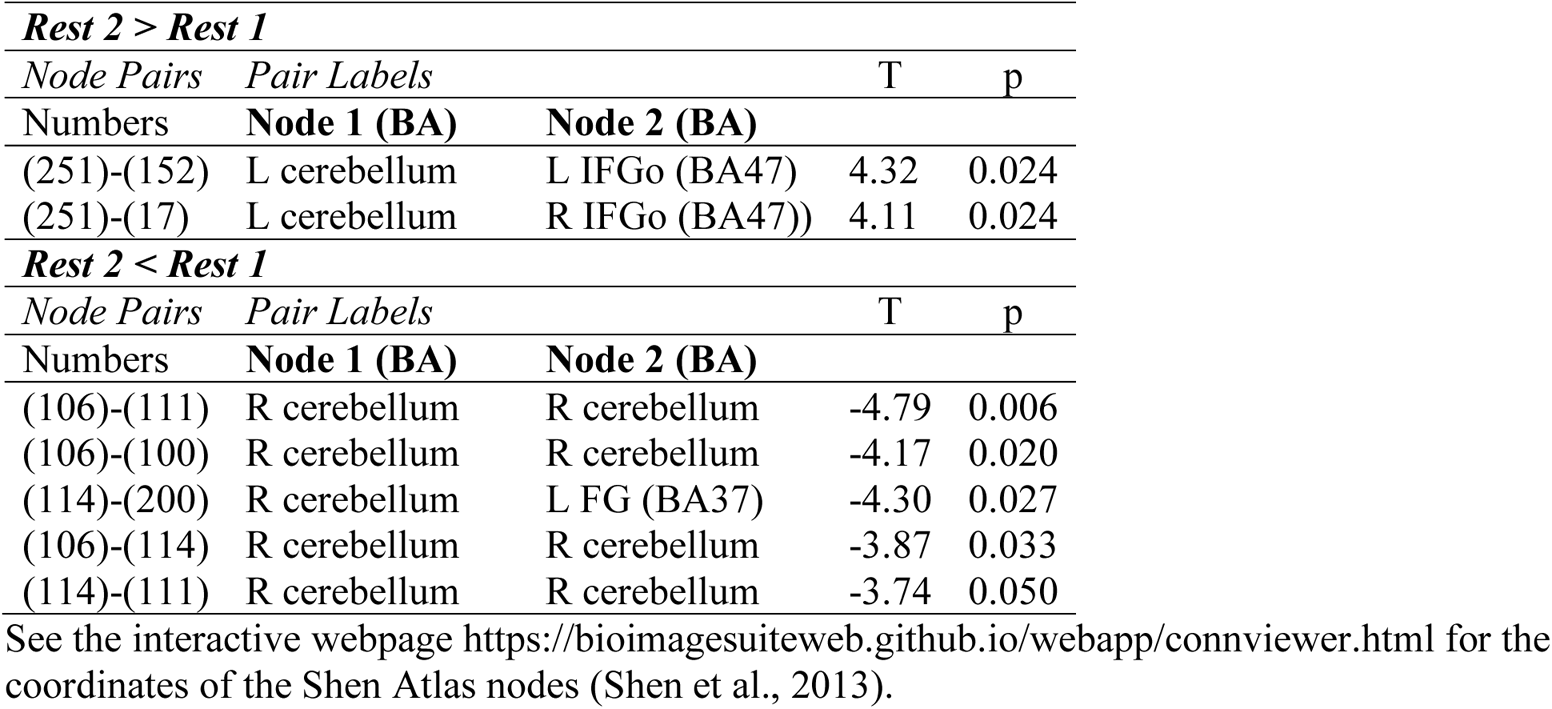
MI-NF > VI resting-state functional connectivity.

| *Rest 2 > Rest 1* |  |  |  |  |
| --- | --- | --- | --- | --- |
| *Node Pairs* | *Pair Labels* |  | *T* | *p* |
| Numbers | **Node 1 (BA)** | **Node 2 (BA)** |  |  |
| (251)-(152) | L cerebellum | L IFGo (BA47) | 4.32 | 0.024 |
| (251)-(17) | L cerebellum | R IFGo (BA47)) | 4.11 | 0.024 |
| *Rest 2 < Rest 1* |  |  |  |  |
| *Node Pairs* | *Pair Labels* |  | *T* | *p* |
| Numbers | **Node 1 (BA)** | **Node 2 (BA)** |  |  |
| (106)-(111) | R cerebellum | R cerebellum | -4.79 | 0.006 |
| (106)-(100) | R cerebellum | R cerebellum | -4.17 | 0.020 |
| (114)-(200) | R cerebellum | L FG (BA37) | -4.30 | 0.027 |
| (106)-(114) | R cerebellum | R cerebellum | -3.87 | 0.033 |
| (114)-(111) | R cerebellum | R cerebellum | -3.74 | 0.050 |

See the interactive webpage https://bioimagesuiteweb.github.io/webapp/connviewer.html for the coordinates of the Shen Atlas nodes (Shen et al., 2013).

**Fig. S3.**
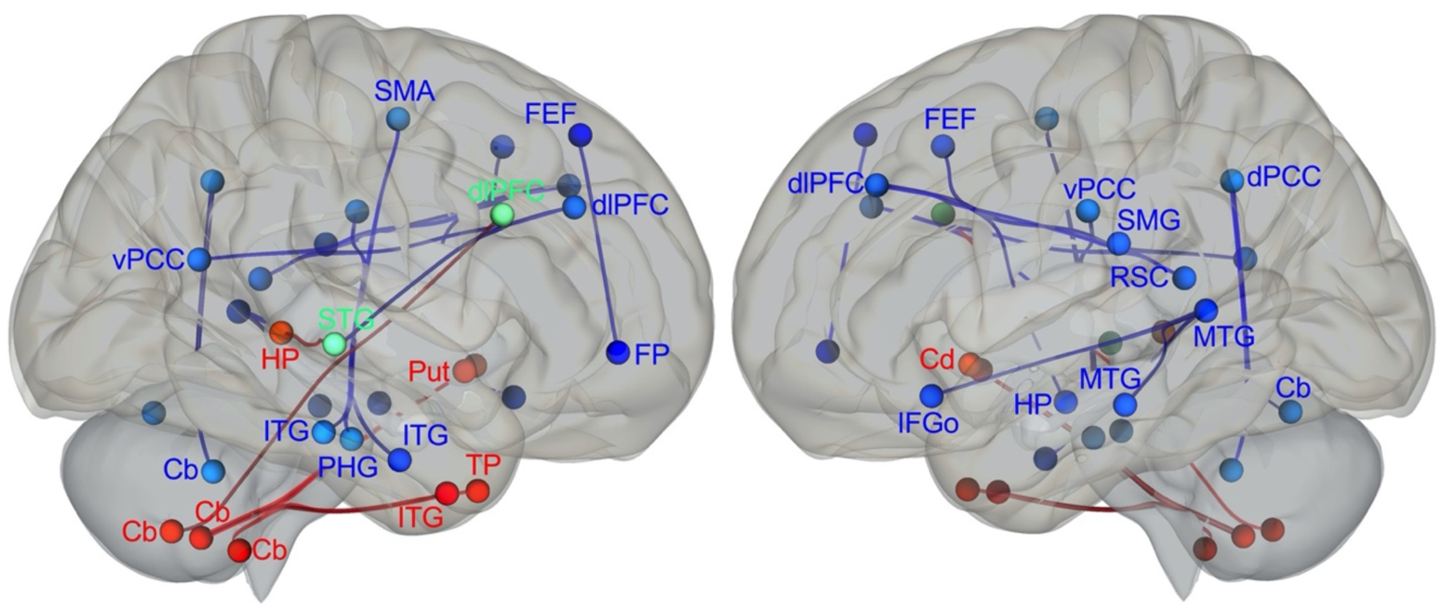
Whole-brain imagery task-based functional connectivity main effect of MI-NF (imagery 2 > imagery 1). Red: increased, blue: decreased. Cb: Cerebellum, Cd: Caudate, dlPFC: Dorsolateral prefrontal cortex, dPCC: Dorsal posterior cingulate cortex, FEF: Frontal eye fields, FP: Frontal pole, HP: Hippocampus, IFGo: Inferior frontal gyrus, orbital part; ITG: Inferior temporal gyrus, MTG: Middle temporal gyrus, PHG: Parahippocampal gyrus, Put: Putamen, RSC: Retrosplenial cortex, SMA: Supplementary motor area, SMG: Supramarginal gyrus, STG: Superior temporal gyrus, TP: Temporal pole, vPCC: Ventral posterior cingulate cortex.

**Table S10.** MI-NF imagery task-based functional connectivity.

| ***Imagery 2 > Imagery 1*** |  |  |  |  |
| --- | --- | --- | --- | --- |
| *Node Pairs* | *Pair Labels* |  | *T* | *p* |
| Numbers | **Node 1 (BA)** | **Node 2 (BA)** |  |  |
| (57)-(115) | R ITG (BA20) | R cerebellum | 5.29 | 0.001 |
| (116)-(22) | R cerebellum | R dIPFC (BA9) | 4.45 | 0.017 |
| (125)-(115) | R putamen | R cerebellum | 4.39 | 0.020 |
| (51)-(118) | R TP (BA38) | R cerebellum) | 4.31 | 0.026 |
| (63)-(93) | R STG (BA22) | R hippocampus | 3.98 | 0.036 |
| (259)-(115) | L caudate | R cerebellum | 4.16 | 0.042 |
| ***Imagery 2 < Imagery 1*** |  |  |  |  |
| *Node Pairs* | *Pair Labels* |  | *T* | *p* |
| Numbers | **Node 1 (BA)** | **Node 2 (BA)** |  |  |
| (193)-(192) | L MTG (BA21) | L MTG (BA21) | -4.93 | 0.004 |
| (231)-(150) | L hippocampus | L FEF (BA8) | -4.81 | 0.005 |
| (56)-(192) | R ITG (BA20) | L MTG (BA21) | -4.48 | 0.015 |
| (12)-(8) | R FEF (BA8) | R FP (BA10) | -4.43 | 0.017 |
| (224)-(59) | L vPCC (BA23) | R ITG (BA20) | -4.38 | 0.021 |
| (96)-(25) | R PHG (BA36) | R SMA (BA6) | -4.27 | 0.029 |
| (146)-(86) | L dIPFC (BA9) | R vPCC (BA23) | -4.23 | 0.033 |
| (192)-(153) | L MTG (BA21) | L IFGo (BA47) | -4.00 | 0.034 |
| (181)-(146) | L SMG (BA40) | L dIPFC (BA9) | -4.21 | 0.035 |
| (63)-(22) | R STG (BA22) | R dIPFC (BA9) | -4.11 | 0.036 |
| (227)-(11) | L RSC (BA30) | R dlPFC (BA9) | -4.14 | 0.043 |
| (225)-(248) | L dPCC (BA31) | L cerebellum | -4.06 | 0.044 |
| (225)-(117) | L dPCC (BA31) | R cerebellum | -3.91 | 0.044 |
| (59)-(224) | R ITG (BA20) | L vPCC (BA23) | -4.13 | 0.045 |

BA: Brodmann area, dlPFC: Dorsolateral prefrontal cortex, dPCC: Dorsal posterior cingulate cortex, FEF: Frontal eye fields, FP: Frontal pole, IFGo: Inferior frontal gyrus, orbital part; ITG: Inferior temporal gyrus, MTG: Middle temporal gyrus, PHG: Parahippocampal gyrus, Put: Putamen, RSC: Retrosplenial cortex, SMA: Supplementary motor area, SMG: Supramarginal gyrus, STG: Superior temporal gyrus, TP: Temporal pole, vPCC: Ventral posterior cingulate cortex. See the interactive webpage https://bioimagesuiteweb.github.io/webapp/connviewer.html for the coordinates of the Shen Atlas nodes (Shen et al., 2013).

**Fig. S4.**
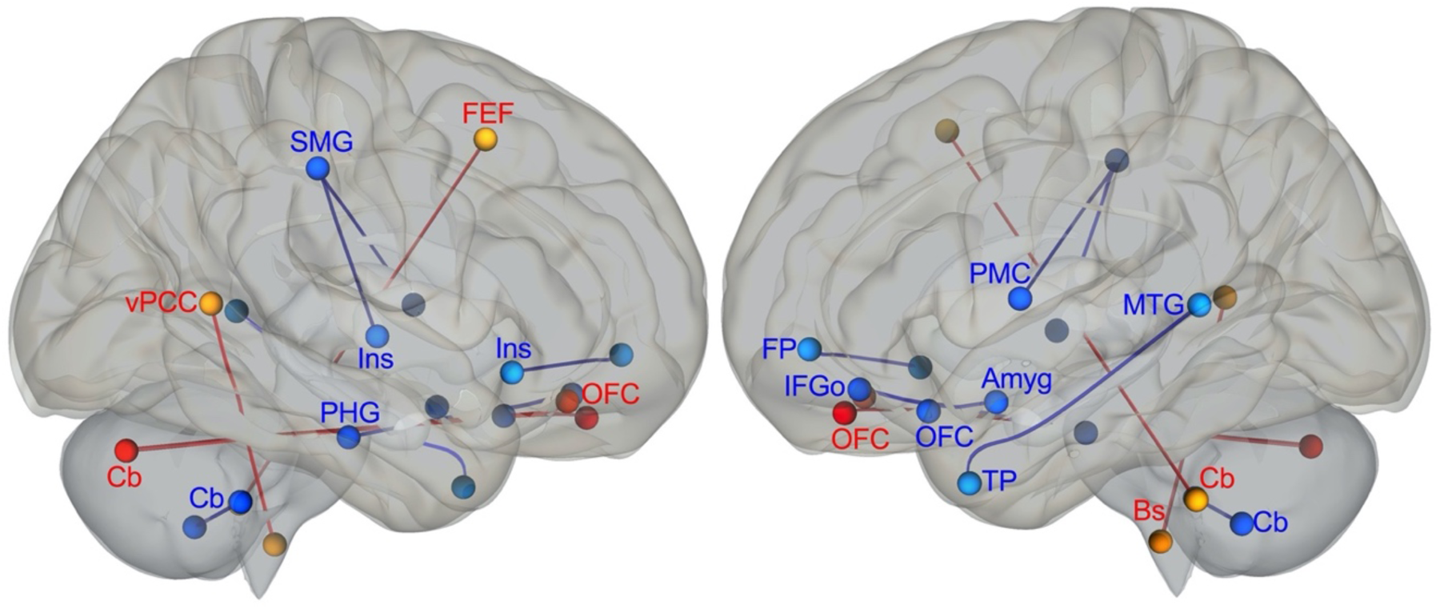
Whole-brain imagery task-based functional connectivity main effect of VI (imagery 2 > imagery 1). Red: Increased, blue: decreased. Amyg: Amygdala, Bs: Brainstem, Cb: Cerebellum, FEF: Frontal eye fields, FP: Frontal pole, IFGo: Inferior frontal gyrus, orbital part; Ins: Insula, MTG: Middle temporal gyrus, OFC: Orbitofrontal cortex, PHG: Parahippocampal gyrus, PMC: Premotor cortex, SMG: Supramarginal gyrus, TP: Temporal pole, vPCC: Ventral posterior cingulate cortex.

**Table S11.** VI imagery task-based functional connectivity.

| ***Imagery 2 > Imagery 1*** |  |  |  |  |
| --- | --- | --- | --- | --- |
| *Node Pairs* | *Pair Labels* |  | *T* | *p* |
| Numbers | **Node 1 (BA)** | **Node 2 (BA)** |  |  |
| (102)-(137) | R cerebellum | L OFC (BA11) | 5.59 | 0.000 |
| (102)-(3) | R cerebellum | R OFC (BA11) | 5.08 | 0.001 |
| (266)-(87) | L brainstem | R vPCC (BA23) | 4.10 | 0.049 |
| (252)-(14) | L cerebellum | R FEF (BA8) | 4.10 | 0.050 |
| ***Imagery 2 < Imagery 1*** |  |  |  |  |
| *Node Pairs* | *Pair Labels* |  | *T* | *p* |
| Numbers | **Node 1 (BA)** | **Node 2 (BA)** |  |  |
| (96)-(152) | R PHG (BA36) | L IFGo (BA47) | -5.22 | 0.001 |
| (45)-(163) | R SMG (BA40) | L PMC (BA6) | -4.55 | 0.012 |
| (250)-(107) | L cerebellum | R cerebellum | -4.29 | 0.027 |
| (45)-(37) | R SMG (BA40) | R insula (BA13) | -3.95 | 0.039 |
| (36)-(138) | R insula (BA13) | L FP (BA10) | -4.17 | 0.040 |
| (192)-(189) | L MTG (BA21) | L TP (BA38) | -4.11 | 0.048 |
| (228)-(135) | L amygdala | L OFC (BA11) | -4.10 | 0.049 |

BA: Brodmann area, FEF: Frontal eye fields, FP: Frontal pole, IFGo: Inferior frontal gyrus, orbital part; MTG: Middle temporal gyrus, OFC: Orbitofrontal cortex, PHG: Parahippocampal gyrus, PMC: Premotor cortex, SMG: Supramarginal gyrus, TP: Temporal pole, vPCC: Ventral posterior cingulate cortex. See the interactive webpage https:// bioimagesuiteweb.github.io/webapp/connviewer.html for the coordinates of the Shen Atlas nodes (Shen et al., 2013).

